# Genomic correlates of clinical CAR-T cell activity

**DOI:** 10.1101/2025.10.08.25337584

**Authors:** Mark B. Leick, Baihe Sun, Filippo Birocchi, Kathleen M.E. Gallagher, Alexandra Bratt, Seunghun Han, Grace Martin, Harrison J. Silva, Rebecca C. Larson, Tyler M. Chinsky, Hoyin Chu, Christopher R. Reilly, Michael C. Kann, Bryan D. Choi, Sabrina Camp, Riaz Gillani, Merle Phillips, Tamina Kienka, Stefanie R. Bailey, Charlotte E. Graham, Max Jan, Nicholas S. Moore, Nora Horick, Justin Budka, Simone Filosto, Chad M. Williams, Ali Hosseini Rad, Rhine R. Shen, Eliezer Van Allen, Saud AlDubayan, Marcela V. Maus

## Abstract

Chimeric antigen receptor (CAR)-T cell therapies demonstrate potent anti-tumor efficacy in hematologic malignancies, yet clinical outcomes remain unpredictable due to the bespoke nature of the treatment, which is manufactured from each patient’s own T-cells. While germline variants are known to influence response to immune checkpoint inhibitors^1, 2^, their role in CAR-T cell therapy is unknown. Here, we pair whole-genome germline sequencing of lymphoma patients from the ZUMA-1^3^ and ZUMA-7^4^ clinical trials of axicabtagene ciloleucel, along with correlative biomarkers and functional assays, to ascertain the impact of germline variants on CAR-T cell behavior. Hypothesizing shared mechanisms of the most common toxicities of CAR-T cells, namely cytokine release syndrome (CRS) with hemophagocytic lymphohistiocytosis (HLH)—a hyperinflammatory syndrome driven by T cell overactivation^5^, we first looked within 17 canonical HLH-associated genes^6^, and identified putative deleterious *STXBP2* variants in 15% of ZUMA-1 patients with toxicity, which were absent in control subjects who did not experience high grade toxicity. Subjects with these variants had elevated baseline IFN-γ and inflammatory cytokines, findings that were recapitulated in engineered *STXBP2*-deficient and *STXBP2*-variant-expressing primary CAR-T cells derived from healthy donors. However, *STXBP2* variant enrichment was absent in ZUMA-7 for this toxicity phenotype, possibly reflecting differences in underlying disease burden and evolving clinical management between the trials. A more expansive genome-wide analysis revealed *ADAMTSL3* (a negative regulator of TGFβ^7^) as the only gene nominally enriched for putative deleterious variants in both ZUMA-1 and ZUMA-7 among control subjects, suggesting a protective effect. Finally, we focused on associations between germline variants and CAR-T cell expansion after infusion, a more objective and granular continuous variable that is strongly associated with clinical response across most CAR-T products^8^. We found a strong association between *PTPN22*, a known negative regulator of T-cell activation ^9–12^ and an autoimmune risk gene^13, 14^, variant status and CAR-T cell expansion in both ZUMA-1 and ZUMA-7, with the patients having the highest level of CAR-T expansion across clinical trials harboring variants in the gene. Together, these findings demonstrate the first clear association between germline variants and the clinical behavior of engineered immune cell therapies, which has implications for cellular therapy design, monitoring, testing, clinical trial design, and patient care.

## INTRODUCTION

Chimeric antigen receptor (CAR) T cell therapy has changed the clinical landscape of hematologic malignancies, with an ever-growing number of FDA approvals since 2017^3,15–20^. Clinical use of CAR T cell products has generally been limited to tertiary academic centers due to the need for close monitoring for two class-effect cytokine-related toxicities, namely cytokine release syndrome (CRS) and immune effector cell-associated neurotoxicity syndrome (ICANS), either of which can be fatal^21^. Unlike traditional therapeutics that are essentially identical across all patients, CAR-T cells are bespoke and harbor all of the ancestral genetic polymorphisms of their parental T-cells. Attempts to predict CAR-T cell behavior have been limited to the use of existing tumor burden or traditional lab parameters, but fail to account for the complexities of these living drugs^22, 23^. In the context of another immunotherapy, immune checkpoint blockade therapy, a detailed survey of immune-related inherited polymorphisms revealed that the gain of a cryptic exon for IL-7, a critical regulator of lymphocyte homeostasis, is strongly associated with immune-related adverse events^1^, demonstrating the importance of the host genetic state in the clinical behavior of immunotherapy.

The biochemical and clinical phenotype of CRS is similar to the rare hyperinflammatory syndrome hemophagocytic lymphohistiocytosis (HLH). Both share a putative mechanism in which highly activated T cells produce cytokines (like IFN-γ) that spur myeloid production of inflammatory mediators (like IL-6, IL-1, and TNFα), which drive systemic toxicities^5, 24^. Among the known etiologies driving HLH are loss-of-function (LOF) germline variants in cytolytic pathways, which display variable penetrance and phenotypic effects^6,25–27^. While initial HLH studies identified only biallelic loss-of-function variants as causative of classical, familial HLH, which typically presents in childhood, a more nuanced understanding now includes heterozygous LOF variants in granule-dependent cytotoxicity gene members, which can result in HLH in the context of significant immunologic challenge^26,28–33^ and/or later in adulthood.

We initially hypothesized that germline variants in HLH-associated genes could modulate CAR-T cell behavior and immune response, resulting in clinically meaningful phenomena. To maximize the ability to explore the full range of variants potentially relevant in this clinical setting, we performed whole genome germline sequencing and correlative analyses with biomarkers and functional assays to study variants in two clinical trials of axicabtagene ciloleucel in patients with aggressive lymphoma, the ZUMA-1^3^ (n=86 evaluable subjects) and ZUMA-7 (n=150 evaluable subjects) clinical trials^4^. We find genomic, correlative cytokine, and corroborative bioassay evidence linking specific germline genomic variants to CAR-T cell behavior in the clinical and experimental settings.

## RESULTS

### Study Participants

A total of 86 patients with aggressive lymphomas who received axicabtagene ciloleucel as part of ZUMA-1^3^ underwent whole genome sequencing (Extended Data Fig. 1-3). Of those patients, 41 reached the combined toxicity endpoint (either high-grade CRS, Neurotoxicity-Events (NE, now known as immune effector cell-associated neurotoxicity syndrome ICANS), or those who received tocilizumab; further details in methods) and were included in the toxicity cohort, while 43 patients had no or reduced toxicity and were considered the control cohort. Two patients from the toxicity cohort and four from the control cohorts were excluded during quality control steps (Methods), leaving 39 patients in each cohort. Consistent with prior studies, inflammatory cytokines related to CAR-T cell toxicity (GM-CSF, IFN-γ, MCP-1, and TNFα) were similar between the two cohorts at baseline; however, peak levels after infusion were significantly elevated in the toxicity cohort, though for some cytokines, the absolute magnitude of the difference was small (**Table 1**). Peak IL-6, which is a known driver of cytokine storm^24^, was significantly higher in the toxicity cohort than the control cohort; however, receipt of IL-6 blockade for CAR-T cell toxicity confounds measurement of this cytokine ^3,5,34,35^. Baseline characteristics, including age, sex, and tumor response rates at 1 month, were similar between the cohorts.

**Table 1.**
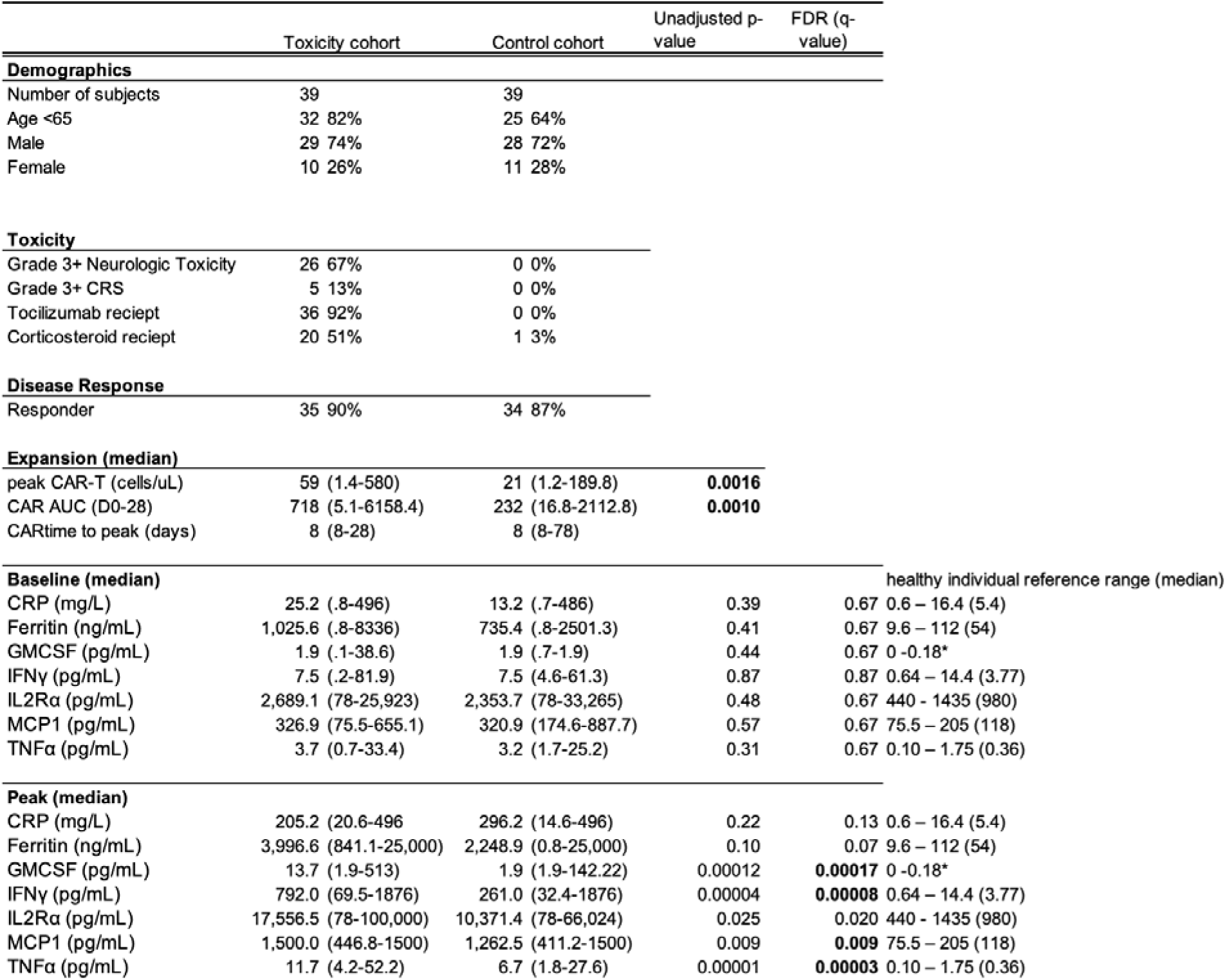
Demographic Characteristics, Clinical Features, and Cytokine Levels of included patients from the ZUMA-1 clinical trial. P-values represent Mann-Whitney U test with a two-stage step-up method of Benjamini, Krieger, and Yekutielei with a false discovery rate (FDR) threshold of 1%. Cytokine values meeting significance threshold (1%) are in bold. *Only detectable in the serum of 5% of healthy subjects.

### Germline Sequencing

Full-depth whole-genome sequencing was performed to explore the full range of variants and improve coverage in coding and adjacent regions^36^. To reduce bias from population structure (i.e., variability in allele frequency patterns across ancestral groups), we limited our genomic analysis to individuals of European ancestry, who also represented the largest subgroup in our cohort. (n=78 of the total n=86, in ZUMA-1).

### Enrichment of germline variants in HLH-associated genes

To determine whether putative deleterious germline variants are associated with toxicity to CAR-T therapy, we systematically characterized germline variants in 17 HLH-associated genes (**Extended Data Table 1**) which were sequenced to a mean coverage of at least 30X (**Extended Data** Fig. 4). For each gene, we compared the burden of putative deleterious germline variants with MAF of <5% (population-level prevalence data were derived from the Genome Aggregation Database [http://gnomad.broadinstitute.org/]) between patients who developed toxicity, (Case Cohort, n=39) vs. ancestry-matched patients who had no significant toxicity (Control Cohort, n=39). Our case-control analysis showed a higher enrichment of putative deleterious germline variants in *STXBP2* in patients who developed toxicity compared to the control cohort. Overall, we identified 6 (15.4%, 95% CI:5.9% to 30.1%) patients who carried heterozygous putative deleterious germline variants in *STXBP2,* all of whom were in the toxicity group, while all patients in the control cohort carried *STXBP2* wild-type alleles (OR>1.44, nominal p value = 0.025, Bonferroni-adjusted p = 0.425 for 17 tests; **Fig. 1a, Extended Data Table 2, Extended Data** Fig. 5). This finding was confirmed through alternative gene burden analysis (SKATO; p=6.52e-3, Bonferroni-adjusted p=0.111 **Extended Data Table 3**). Because the administration of tocilizumab involves clinical subjectivity, we performed a secondary gene burden analysis where we excluded patients whose sole inclusion criteria into the toxicity cohort was receipt of tocilizumab. After excluding tocilizumab from the combined toxicity endpoint criteria, we find an enhanced signal with a stronger association than the primary burden analysis (with tocilizumab: p=6.52e-3, Bonferroni-adjusted p=0.111; excluding tocilizumab: p=1.06e-3, Bonferroni-adjusted p=0.0180) Extended Data Fig. 6). Furthermore, when comparing toxicity-control cohort enrichment of rare putative deleterious variants versus all variants, we find significant enrichment only of the former, suggesting some degree of negative selection (rare putative deleterious variants: OR=inf, 95% CI: 1.44-inf, p=0.025; all *STXBP2* variants: OR=0.443, 95% CI: 0.121-1.615, p=.0347). Notably, neither cases nor controls carried truncating variants in the previously reported HLH-associated genes. All putative deleterious variants were rare missense substitutions, half of which have previously been shown to impact the protein function. A summary of discovered germline variants across the HLH-associated genes can be found in **Extended Data Table 4**.

**Figure 1.**
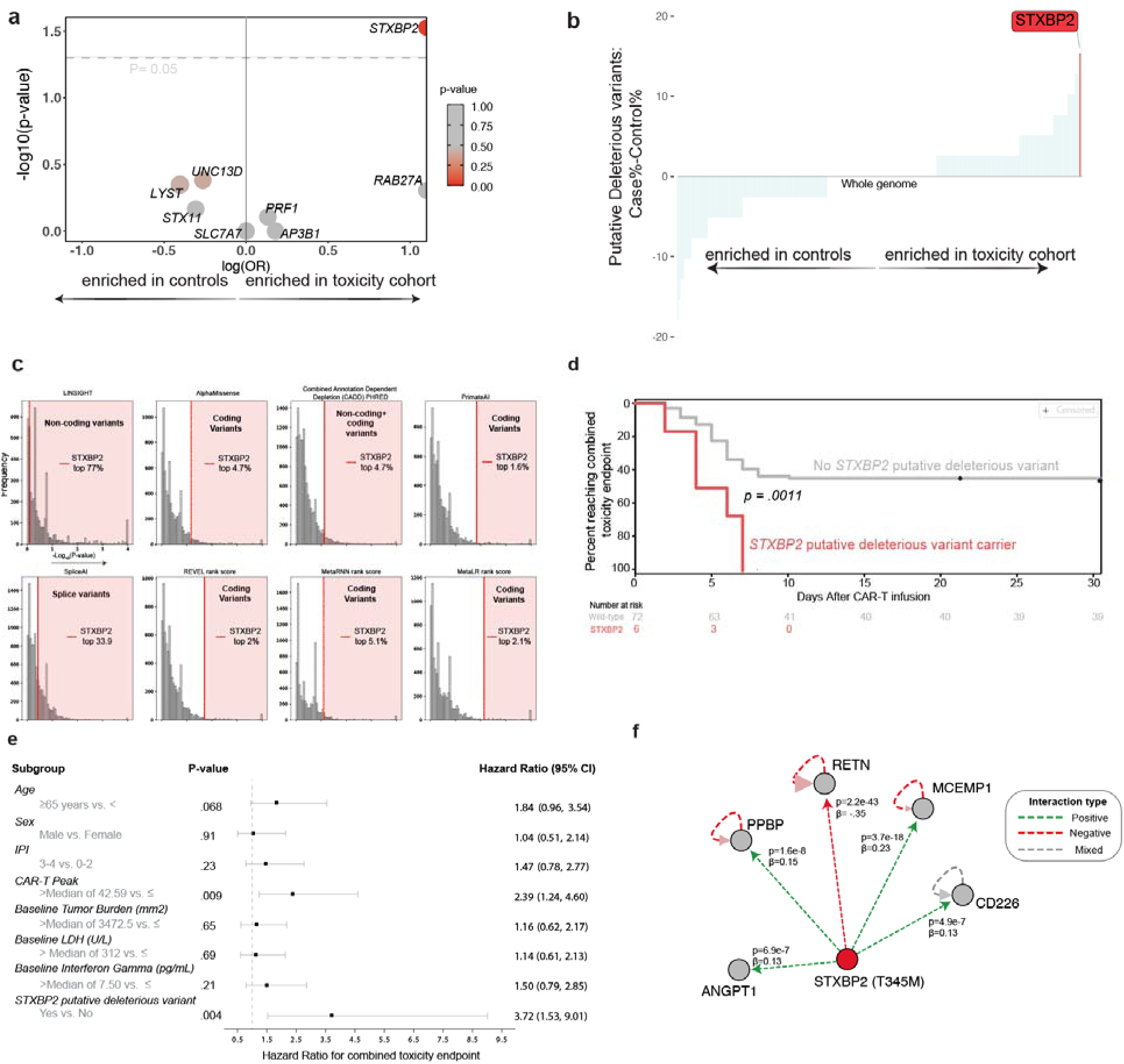
Putative deleterious variants in *STXBP2* are enriched in the toxicity cohort of axicabtagene ciloleucel-treated patients **a**. Gene burden-based variant stratification in HLH-associated genes among ancestry-matched EUR patients. P-value represents Fisher’s Exact test (unadjusted). Only genes in which putative deleterious variants had a non-zero odds ratio are shown. Half circles represent odds ratio (OR) outside graph limits (see **table S2**) **b.** Putative deleterious variant abundance across the genome. The percent difference in putative deleterious abundance in the toxicity cohort minus control cohort (y-axis) for all genes with a putative deleterious variant (n=14,415) in our cohort is displayed in ascending order across the entire genome (x-axis). **c.** Genome-wide distribution of –log_10_(p-values) from analysis of differential deleteriousness across various impact prediction tools. “Rank score” indicates that deleteriousness scores were normalized by rank across all genes. P-value calculated by Mann-Whitney U test. Higher –log_10_(p-values) indicate stronger statistical evidence for differences in predicted deleteriousness of variants found in a given gene between the control and toxicity cohorts. *STXBP2* is ranked among the top ∼5% of genes by differential deleteriousness in impact predictions assessing coding variants, but not those assessing non-coding or splice variants. **d.** Event curve displaying attainment of the combined toxicity endpoint for *STXBP2* putative deleterious variant carriers versus the remainder of the cohort, including only EUR patients. P-value calculated by Log-rank (Mantel-Cox) test. **e**. Hazard ratios for selected clinical and demographic variables. Time from initial treatment to a combined toxicity event or to being censored greater than 30 days is truncated at 30 days. Hazard ratios and 95% CIs are calculated from a Cox proportional hazard model. IPI-international prognostic index, LDH-lactate dehydrogenase, **f.** CGR Proteogenomics Portal^48^ result for *STXBP2,* including all types of association and only protein-truncated and missense/inframe variants. Only primary impacts are shown.

To assess whether this enrichment was driven by systematic sequencing differences between cases and controls, we used the same case-control analysis framework to analyze synonymous as well as common missense variants, two classes of germline variants that are not expected to impact protein function. The prevalence of synonymous and common missense variants in *STXBP2* was similar in toxicity vs. non-toxicity groups (adjusted p-value p=1 for each comparison), corroborating that the identification of putative deleterious variants was not due to systematic sequencing issues (**Extended Data Table 5-6, Extended Data** Fig. 7).

To ascertain if *STXBP2* putative deleterious variants were enriched in lymphoma patients in general compared to healthy control subjects, we looked at the prevalence of *STXBP2* putative deleterious variants in a control cohort of 19,757 ancestry-matched individuals without known malignancy. We found no significant enrichment of *STXBP2* putative deleterious variants in our lymphoma cohort relative to healthy controls (7.69% in this population, and 7.63% in the general population, p = .99; Extended Data Fig. 8).

### Germline variants discovered in *STXBP2*

A total of 6 patients in the toxicity cohort carried a putative deleterious variant in *STXBP2* (Extended Data Fig. 9). *STXBP2* putative deleterious variants had a population-level minor allele frequency in the range of 0.002% to 1.2% as reported by the Genome Aggregation Database (gnomAD, https://gnomad.broadinstitute.org/). Two of the four variants (p.Thr345Met [c.1034C>T], p.Ala433Val [c.1298C>T]), representing 4 of 6 patients with *STXBP2* variants in our cohort, have been previously identified in a heterozygous fashion in patients with HLH (**Extended Data Table 7**) and confirmed to mechanistically disrupt degranulation *in vitro^31,37–43^*. An HLH patient with a variant (p.Arg575_Phe576del) at the same residue as one of the remaining two variants (p.Arg575Cys [c.1723C>T;]) in our cohort had also been previously reported, supporting the potential importance of this region for protein function^44^. The remaining variant, p.Arg474Cys [c.1420C>T], lies within a highly conserved region of protein family members and is adjacent to the previously established canonical driver of familial hemophagocytic lymphohistiocytosis type 5 (FHL-5), p.P477L, suggesting that protein alterations in this highly conserved region are poorly tolerated (Extended Data Fig. 10)^45^. Furthermore, *in silico* pathogenicity predictors and structural modeling were predictive of deleterious effects on protein function (Extended Data Fig. 11).

While our analysis began with the *a priori* hypothesis of an association between variants in HLH-related genes and CRS, given how divergent *STXBP2* putative deleterious variant stratification was compared to other HLH-associated variants, we performed an exploratory analysis across all genes. A difference in putative deleterious variants between the toxicity cohort and controls that was as divergent as *STXBP2* could be found in less than 1% of other genes in the entire genome (**Fig. 1b**). Furthermore, *STXBP2* shows one of the most significant differences in predicted variant deleteriousness between control and toxicity cohorts, found to be in the top 5% of genes ranked by differential deleteriousness genome-wide across multiple pathogenicity prediction tools (**Fig. 1c**).

Like the majority of genes implicated in HLH^46^, *STXBP2* is involved in lymphocyte degranulation, and homozygous loss-of-function variants in the gene define familial hemophagocytic lymphohistiocytosis type 5 (FHL-5)^45^, though heterozygous cases have been identified as well^31–33,37,38,40,43^. *STXBP2* plays an important role in intracellular trafficking, control of soluble NSF attachment protein receptor (SNARE) complex assembly, and release of granules mediating cytotoxicity (Extended Data Fig. 12). Germline variants in *STXBP2* resulting in familial HLH have been identified across imputed protein domains.

Similarly, those identified in this study were not restricted to any one region, and *STXBP2* variants (including p.A433V identified in this cohort) have been demonstrated to act in a dominant negative fashion^31,32,47^.

### Clinical and biochemical profiles of patients with *STXBP2* putative deleterious variants

To explore the clinical profile of patients with putative deleterious variants in *STXBP2*, we analyzed the time at which patients developed significant toxicity (as defined by the combined toxicity endpoint). By day 10 after CAR-T infusion, all patients with *STXBP2* putative deleterious variants reached the combined toxicity endpoint (though with similar rates of high-grade neurologic toxicity 67% vs 72%, and CRS 17% vs 13% to the broader study), while only half of the remaining total patients met any of the toxicity endpoints (100% vs. 50%, p=0.0015) (**Fig. 1d**). Moreover, *STXBP2* putative deleterious variant carriers reached the combined toxicity endpoint more quickly, at a median of 5 days after CAR-T infusion compared to 7 days for those non-carriers who reached the combined toxicity endpoint. Furthermore, when patients whose sole criterion of the toxicity endpoint was receipt of tocilizumab were removed, we found a trend towards earlier development of high-grade CRS or neurologic toxicity among *STXBP2* variant carriers (p=0.1, Extended Data Fig. 13). This was true for both the CRS and neurotoxicity components of the combined toxicity endpoint, with median day of high-grade CRS onset (4 vs. 6.5 days) and neurotoxicity (6 vs. 7 days) for variant carriers versus the remaining toxicity cohort, respectively.

In a multivariate analysis, the presence of an *STXBP2* putative deleterious variant was more strongly predictive (hazard ratio 3.72, 95% confidence interval 1.53 to 9.01) of toxicity than other established predictors of CAR-T cell toxicity, such as baseline tumor burden (hazard ratio 1.16, 95% confidence interval 0.62 to 2.17) or CAR-T cell peak expansion (hazard ratio 2.39, 95% confidence interval 1.24 to 4.60) **(Fig. 1e**)^3^. *STXBP2* putative deleterious variant carriers were distributed across sex and had similar clinical tumor response rates to the remainder of ZUMA-1 patients (**Extended Data Table 8**). Leveraging a recent study that drew rare variant associations between 49,736 subjects in the UKBiobank with the quantification of 2,923 plasma proteins from each subject ^48^, we found that one of our variants, STXBP2^T345M^, was strongly associated with perturbations in multiple serum proteins thought to be involved in inflammatory pathways, including ANGPT1, PPBP, RETN, MCEMP1, and CD226.^49–51^ (**Fig. 1f**). CAR-T peak expansion and area under the curve (AUC) of *STXBP2* putative deleterious variant carriers were overall comparable to cohort medians, however, both p.A433V (which has previously been shown to have a dominant negative effect^31^) carriers had pharmacokinetic values that exceeded the toxicity cohort median (peak: 75 and 78 vs 59 for cohort median; AUC: 754 and 934 vs 733 for cohort median respectively). Notably, these two patients developed high-grade neurotoxicity, which could suggest some differences in clinical severity and phenotype among our identified putative deleterious *STXBP2* variants. Both progression-free survival and duration of response were similar between variant carriers and non-carriers (Extended Data Fig. 14). Variant carriers also had similar levels of baseline tumor burden, and the tumor marker lactate dehydrogenase (LDH) and received their first dose of tocilizumab a similar number of days after CAR-T infusion and total number of doses as the remainder of the toxicity cohort (Extended Data Fig. 15). Prolonged cytopenias (grade 3 or 4 neutropenia or thrombocytopenia persisting beyond 30 days) were seen in 2/6 (33%) patients with *STXBP2* putative deleterious variants versus 21/72 (29%) of the entire cohort and 11/33 (33%) of the toxicity cohort. One patient (*STXBP2* wild type) in ZUMA-1 met diagnostic criteria for HLH.^52^

To further investigate the biochemical consequences of putative deleterious *STXBP2* variant carrier status in response to axicabtagene ciloleucel treatment in patients, we stratified pre-treatment and post-infusion cytokines by variant status. Patients harboring *STXBP2* putative deleterious germline variants had higher levels of IFN-γ at baseline, prior to CAR-T infusion, than the remainder of the toxicity cohort (median 30.5 vs. 7.5 pg/mL, respectively; p=.022, **Fig. 2a**), as well as all other non-variant carriers irrespective of toxicity status (median 30.5 vs. 7.5 pg/mL; p= .0086). After infusion, peak levels of the inflammatory cytokines IFN-γ, IL2Rα, and TNFα were collectively higher (p=.032) among *STXBP2* putative deleterious variant carriers than *STXBP2* wild-type patients. Furthermore, among HLH-associated genes, possession of putative deleterious variants in *STXBP2* was significantly associated with higher IFN-γ levels across the cohort (p= 0.0164; Extended Data Fig. 16)

**Figure 2.**
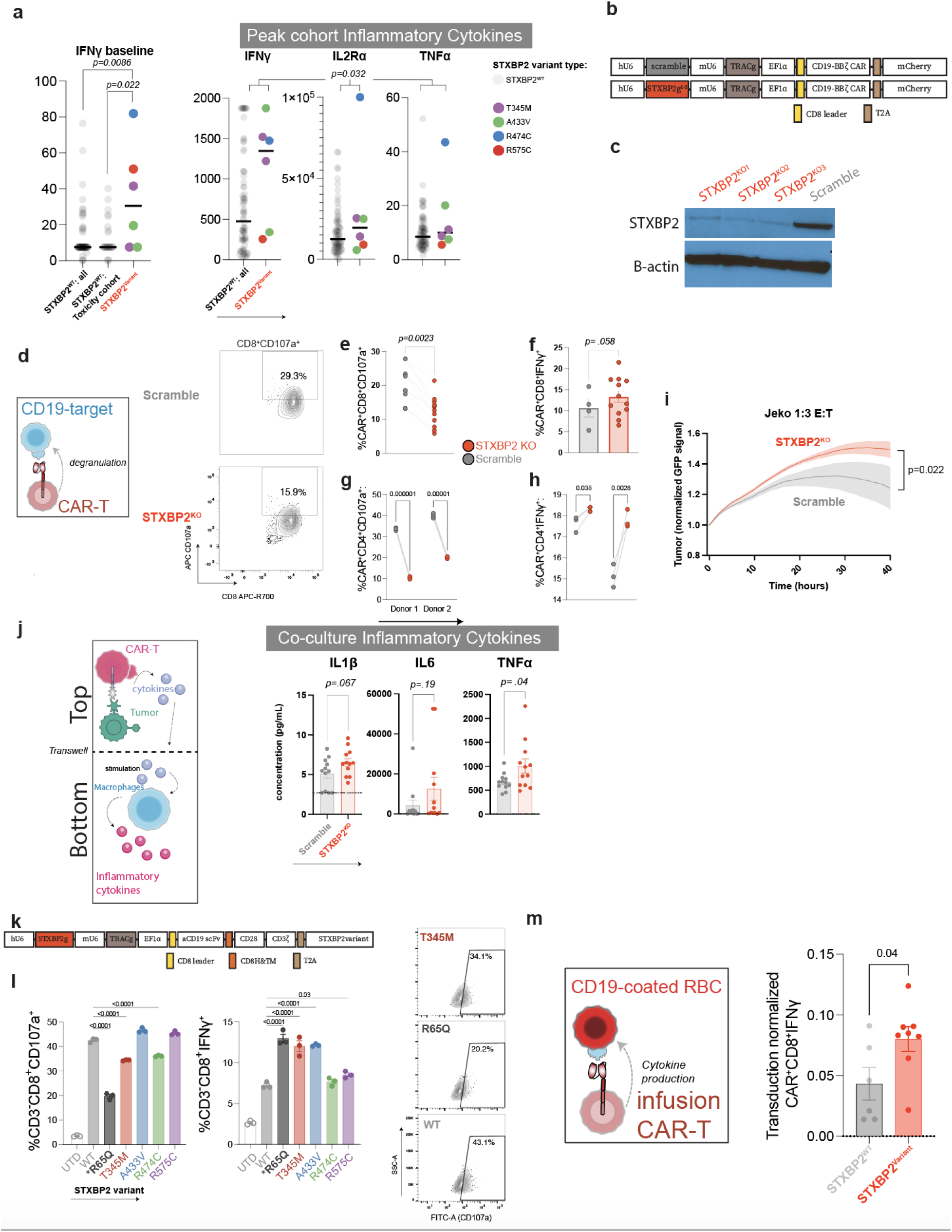
*STXBP2* loss is associated with inflammatory cytokine production in patients and *in vitro* **a.** Inflammatory cytokine comparison of *STXBP2* putative deleterious variant carriers compared to the remainder of the toxicity cohort or entire cohort. IFN-_ baseline p-values represent two-tailed Mann-Whitney U tests, peak cytokine p-value represents two-tailed vanElteren (stratified Wilcoxon rank sum) test. For ZUMA-1: IFN_: lower limit of quantification (LLOQ) is 7.5 pg/ml TNFa: LLOQ is 0.7 pg/ml; for ZUMA-7 IFN_: LLOQ is 1.43 pg/ml TNFa: LLOQ is 0.3 pg/ml **b.** _CD19-CD28 *STXBP2* knockout and scrambled guide control CAR-T designed by Maus lab investigators and does not represent the exact structure of axicabtagene ciloleucel **c**. Western blot of _CD19-CD28 *STXBP2* knockout and scrambled guide control CAR-T cells in primary human T cells (full scan of the western blot is available in Extended Data Fig. 26). **d.** Co-culture schematic of _CD19-CD28 CAR-T cell degranulation assay and representative flow cytometric plots. CAR-T cells were co-cultured with CD19-expressing K562 targets for 4 hours, then stained for CD107a and intracellular IFN_. **e.** CAR-T cell degranulation (CD8+CD107a+) shown for three distinct CRISPR-Cas9 *STXBP2* knockout guides and one scrambled guide control construct. Data points inclusive of two separate experimental replicates with three unique CRISPR guides and CAR-T cells from two donors. P-value represents two-tailed ratio paired t-test. **f**. CAR-T cell intracellular IFN-_ after 4-hour co-culture with CD19-expressing K562 target cells. Data points inclusive of two separate experimental replicates with three unique CRISPR guides and CAR-T cells from two donors. P-value represents one-tailed ratio paired t test. **g.** Primary STXBP2 KO CAR-T cells were generated using just one of the three *STXBP2* CRISPR-Cas9 guides (or scramble guide control) from c-f and were then co-cultured with Nalm6 leukemia targets for 4 hours and CD107a and intracellular IFN-_ (**h.**) were evaluated by flow cytometry. p-values by unpaired t-test. **i**. Real time cytotoxicity assay comparing *STXBP2* knockout and scrambled guide control CAR-T cells against Jeko lymphoma target cells (see **Extended Data Fig. S17** for additional targets). Data points include three unique CRISPR guides and CAR-T cells from two donors. Mean± SEM is shown. P-value represents 2-way ANOVA. **j.** Experimental schematic: *STXBP2* knockout or scramble control CAR-T cells were co-cultured with Nalm-6 tumor targets separated by a transwell from GM-CSF polarized macrophages. Supernatant was collected after 72 hours and analyzed for cytokines. Dotted lines represent the lower limit of detection. P-values represent two-tailed unpaired t-test representing 3 unique CRISPR knockout guides and n=4 donors (two T cell donors and two macrophage donors). **k.** Schematic for *STXBP2* knockout and variant overexpression constructs. **l.** Primary T-cells transduced with the constructs in k. were expanded and selected for edited T-cells prior to use in a 4-hour co-culture with Nalm6 leukemia target cells. Cells then underwent flow cytometry for the indicated markers with representative flow plots. See Extended Data Fig. 18 and 19 for additional results. Bars represent the mean of technical triplicates. P-values represent 2way ANOVA. **m.** CAR-T cells from the infusion product of ZUMA-1 *STXBP2* variant carriers, or control non-variant carriers, were stimulated by CD19-coated red blood cells for four hours followed by assessment by flow cytometry. Two additional patients from ZUMA-1 cohort 3 with *STXBP2* variants were included (excluded from the genomic analysis due to prophylactic receipt of tocilizumab) IFNγ production was normalized to CAR-T cell expression. Bars represent the mean±SEM and p-values are unpaired t-tests. See Extended Data Fig. 20 for additional results.

### Consequences of *STXBP2* loss in CAR-T cells

We then sought to model the effects of *STXBP2* deficiency in CAR-T cells engineered from healthy donors. We generated αCD19-CD28 CAR-T cells (similar in design to the axicabtagene ciloleucel transgene), in which CRISPR-Cas9 was utilized to knock out *STXBP2* (**Fig. 2b and 2c**) in primary healthy donor T cells. In an experiment modeled on the NK-degranulation assay used for clinical diagnosis of HLH^53^, we co-cultured the CAR-T cells with CD19-expressing K562 stimulator cells and assessed short-term degranulation by measuring levels of lysosome-associated membrane protein-1 (LAMP-1/ CD107a), which represents turnover of the interior cytotoxic granules in lymphocytes.

*STXBP2*-KO CAR T cells demonstrated significantly impaired degranulation, impaired cytotoxicity (CD8^+^=*p=.0023,* CD4^+^ p=<.0001), and increased intracellular IFN-γ production (CD4^+^ p=.0028, but not strictly statistically significant for CD8^+^ cells *p=.058*, **Fig. 2d-i, Extended Data** Fig. 17).

Current models of CRS hypothesize that CAR-T cell activation leads to the secretion of mediators like IFN-γ, which in turn activate macrophages to produce inflammatory cytokines like IL-6, IL-1, and TNFα that mediate hyperpyrexia, hypoxia, and capillary leak^24^. To model this phenomenon *in vitro,* we performed a co-culture with our αCD19-CD28*^STXBP2^*^KO^ CAR-T cells and tumor cells separated from macrophages by a transwell membrane, which allows cytokines to pass through (**Fig. 2j**). Analysis of supernatants from the co-cultures revealed higher levels of inflammatory cytokines like IL-6 and TNFα, which mirrored those induced in the serum among *STXBP2* putative deleterious variant carriers in the clinical cohort.

To more closely model the behavior of our specifically identified *STXBP2* variants, we also generated primary human CAR T cells with the same *STXBP2* knockout and αCD19-CD28 transgene, but now also engineered to express each of the variant forms of the STXBP2 protein, including a known dominant negative version, STXBP2^R65Q, 47^ (**Fig. 2k**). When these *STXBP2-*variant expressing CAR-T cells were co-cultured with wild-type or death receptor-deficient leukemia targets, they demonstrated impaired degranulation and elevated intracellular IFN-γ levels, with differences most pronounced in CD8^+^ rather than CD4^+^ T cells and with wild-type rather than death receptor deficient leukemia targets (**Fig. 2l, Extended Data** Fig. 18**, 19**). Finally, ZUMA-1 STXBP2^WT^ subjects, and ZUMA-1 STXBP2^variant^ carrier CAR-T cells were stimulated in a short-term co-culture with CD19-coated red blood cells, which lack extraneous, potentially confounding surface molecules. We found significantly enhanced CD8^+^ IFN-γ production and numerically diminished degranulation, mirroring our findings in the STXBP2 knockout and variant overexpression experiments (**Fig. 2m, Extended Data** Fig. 20).

### Genomic Characterization of the ZUMA-7 Cohort

To enhance study power and explore potential determinants of toxicity in CAT-T treated lymphoma patients beyond the predefined HLH genes, we next performed full-depth whole genome germline sequencing of 150 patients treated in the ZUMA-7 randomized clinical trial of axicabtagene ciloleucel for patients with diffuse large B-cell lymphoma, finding 115 of European ancestry with 71 meeting the same criteria for the toxicity cohort and 44 in the control cohort (**Table 2**, Extended Data Fig. 21). Importantly there were key differences in the patient populations between the ZUMA-1 and ZUMA-7 trials, the number of prior therapies (ZUMA-1 69%≥three prior lines of therapy vs. ZUMA-7 where all subjects had 1 prior therapy) including different levels of CAR-T expansion in the toxicity (but not control) cohorts of both groups (Extended Data Fig. 22) and evidence of significantly higher levels of baseline inflammation (ferritin, p=0.000008) in subjects in ZUMA-1 relative to ZUMA-7. With these differences in mind, we did not identify the same enrichment of *STXBP2* putative deleterious variants among the toxicity cohort in ZUMA-7 (Extended Data Fig. 23).

**Table 2.**
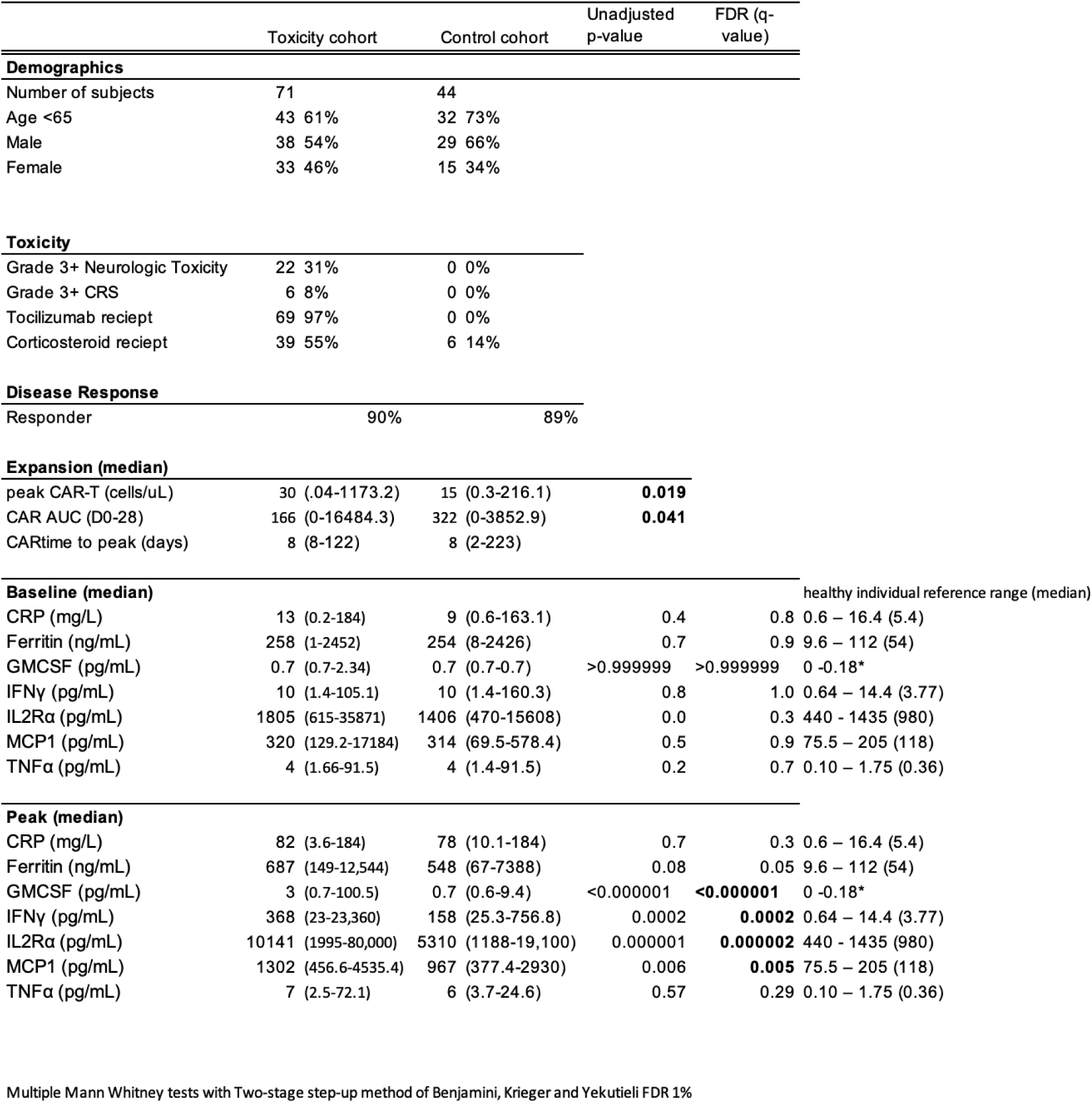
Demographic Characteristics, Clinical Features, and Cytokine Levels of included patients from the ZUMA-7 clinical trial. P-values represent Mann-Whitney U test with a two-stage step-up method of Benjamini, Krieger, and Yekutielei with a false discovery rate (FDR) threshold of 1%. Cytokine values meeting the significance threshold (1%) are in bold. *Only detectable in the serum of 5% of healthy subjects. AUC-area under the curve, the integral of CAR-T cells calculated over a 28-day period from infusion.

We used the availability of these two highly annotated whole genome datasets to explore if there were any genes enriched for putative deleterious variants among the toxicity or control cohorts across the genome in both studies (**Fig. 3a**). One gene, *ADAMTSL3*, achieved nominal (non-FDR corrected) significance in the control cohorts across both the ZUMA-1 and ZUMA-7 studies (**Fig. 3b**). Interestingly, *ADAMTSL3* is a known negative regulator of TGFβ signaling, suggesting a potential protective mechanism of CAR-T cell mediated toxicity ^7^. ZUMA-1*^ADAMTSL3^*variant carriers had significantly lower peak levels of IL-15 (p=.006), with a similar trend among ZUMA-7*^ADAMTSL3^*variant carriers (p=0.19, **Fig. 3c**). Blood and CSF levels of IL-15 have been strongly correlated with neurologic toxicity and CRS across studies, including in ZUMA-1^54–60^. ZUMA-7^ADAMTSL3^ variant carriers also had a lower level of the T-cell recruitment chemokine, CXCL10 (p=.058, difference not evaluable in ZUMA1, Extended Data Figure 24).

**Figure 3.**
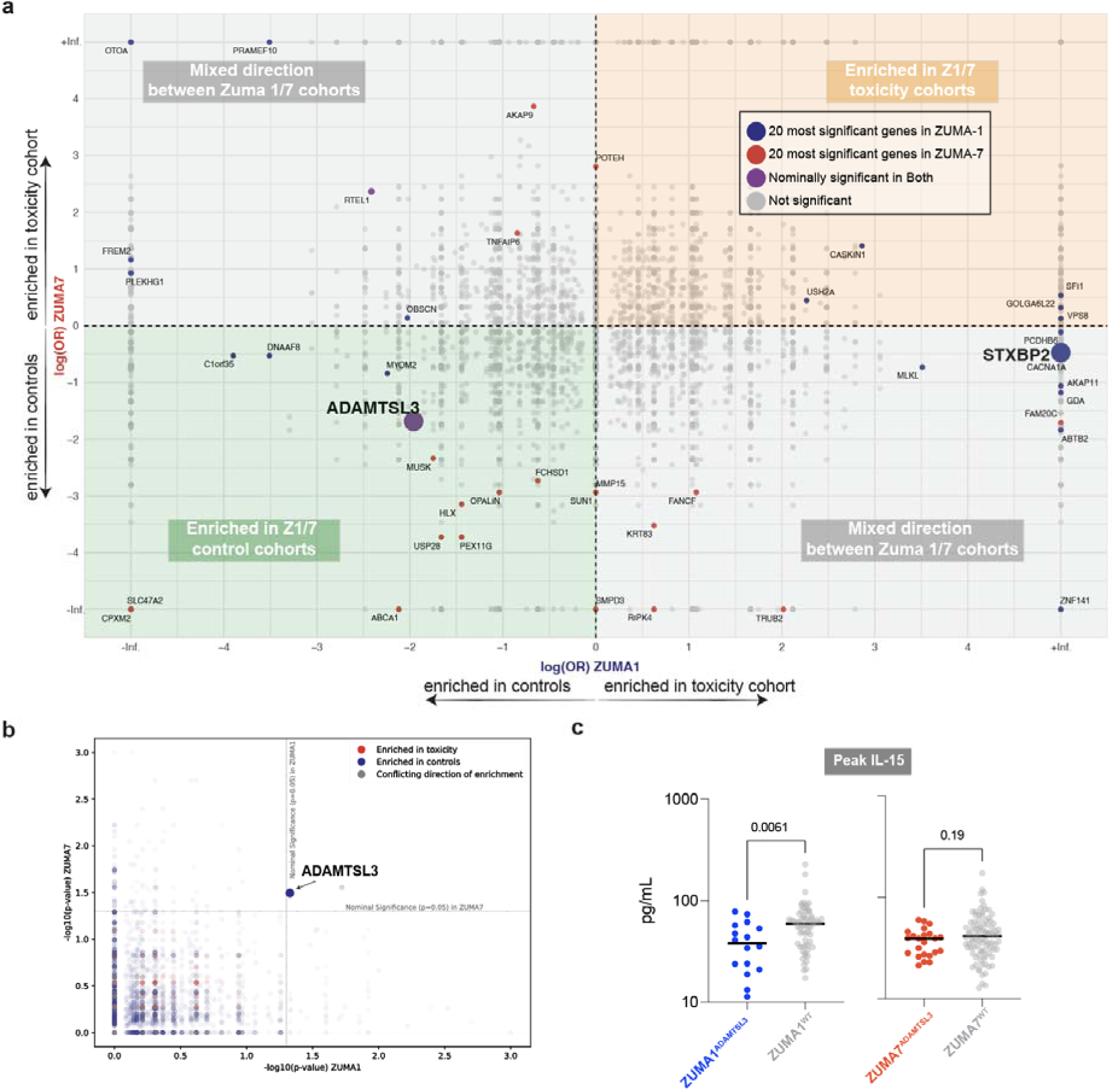
Whole genome association with combined toxicity endpoint in the ZUMA-1 and ZUMA-7 clinical trials. **a.** Genome-wide gene-level enrichment of putative deleterious variants across toxicity and control patients in ZUMA-1 and ZUMA-7. Results plotted by log_2_(OR) for ZUMA-1 and ZUMA-7. The top 20 genes ranked by p-value are labelled and colored corresponding to significance (p<0.05) in ZUMA-1 and ZUMA-7. P-value by Fisher’s Exact test**. b.** Genome-wide gene-level enrichment of putative deleterious variants across toxicity and control patients in ZUMA-1 and ZUMA-7. Results plotted by -log_10_(p-value) for ZUMA-1 and ZUMA-7 with nominal significance (p=0.05) cutoffs as shown. P-value by Fisher’s Exact test. Colors denote enrichment in toxicity or controls, where conflicting directions of enrichment indicate direction of enrichment was inconsistent between ZUMA-1 and ZUMA-7. *ADAMTSL3* is highlighted as the only gene to be nominally significant in both cohorts with the same direction of enrichment. **c.** Level of IL-15 among ZUMA-1 and ZUMA-7 *ADAMTSL3* variant carriers versus non-carriers. P-value by Mann-Whitney U test.

We then hypothesized that although our cohorts are only modestly sized to uncover highly significant gene burden enrichment across the genome, and recognizing the complexity inherent in using toxicity grading as a variable, we turned instead to correlating germline variants with the objective, continuous, and quantitative variable of CAR-T cell expansion, which generally strongly correlates with response across CAR-T cell products ^8^. One gene, *PTPN22*, exceeded Bonferroni significance levels across the genome in both cohorts (ZUMA1: p = 6.18 × 10^-9^, FDR = 8.16 × 10^-6^, Bonferroni-adjusted q = 4.59 × 10^-5^; ZUMA7: p = 7.16 × 10^-15^, FDR = 1.16 × 10^-12^, Bonferroni-adjusted q = 5.31 × 10^-11^; **Fig. 4a**). Variant carriers spanned both the toxicity and control cohorts of both studies and were distributed throughout the protein (**Extended Data figure 25**). PTPN22 is a negative regulator of the T-cell receptor (TCR), dephosphorylating LCK and ZAP70 after TCR activation, and variants in the gene are closely associated with multiple autoimmune conditions^14^. *PTPN22* is related to *PTPN2*, another phosphatase that is a negative regulator of T-cell function, and which is a recurrent top hit in multiple CRISPR knockout screens seeking to augment T-cell function^61, 62^. *PTPN22* variant carriers had the highest expansion of CAR-T cells in both ZUMA-1 and ZUMA-7, despite making up just 5% of the subjects in both cohorts (**Fig. 4b**), though there appears to be a spectrum of penetrance with respect to effect on CAR-T cell expansion within our cohort (**Fig. 4c**). Two of the three highest expanders in ZUMA-1 possessed the K750N variant which lies immediately adjacent to a critical phosphorylation site for PTPN22 (Ser751) function to inhibit proximal TCR signaling and consistently scored higher than almost all of the other variants across pathogenicity predictors^9^.

**Figure 4.**
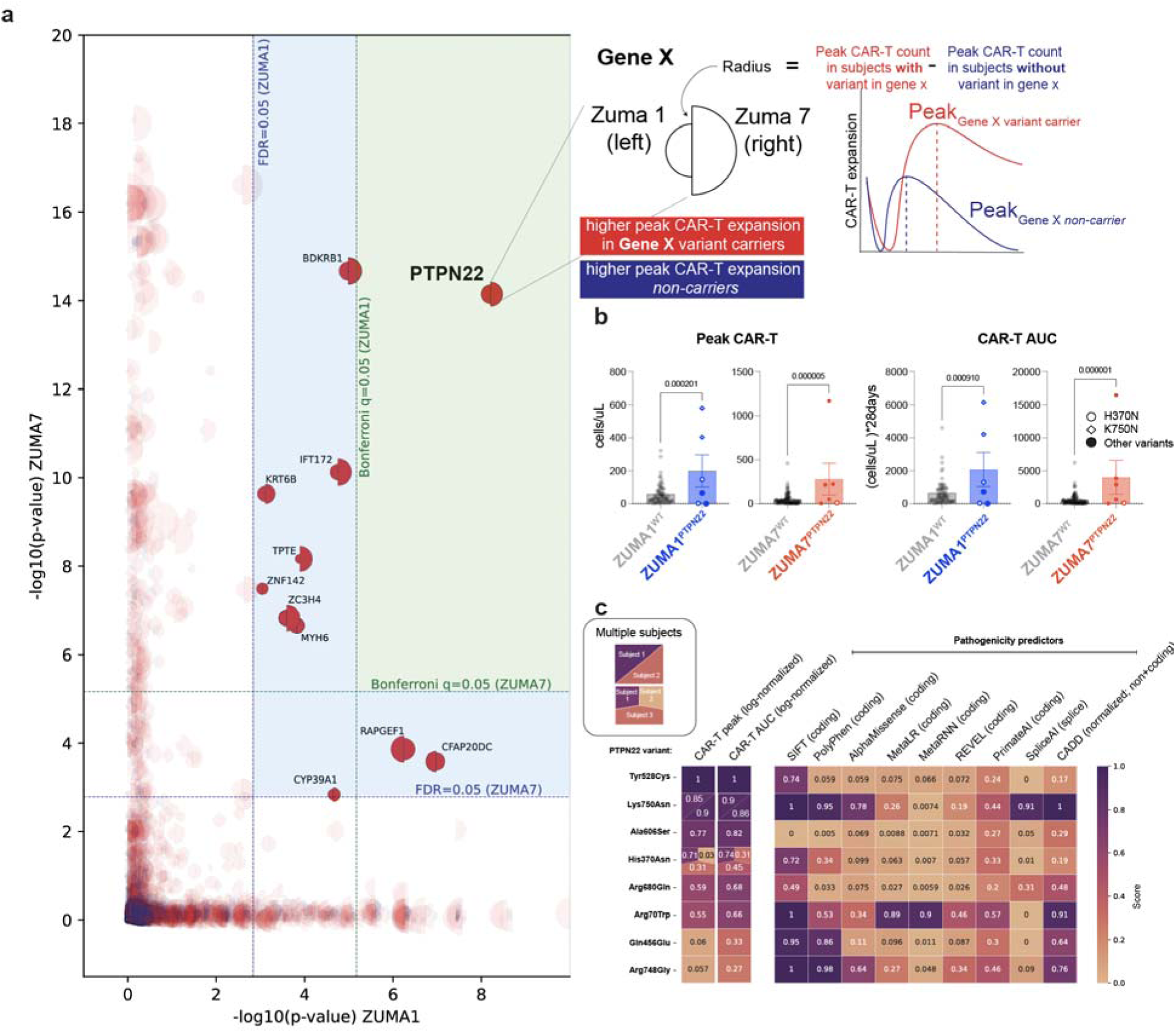
Unbiased association between CAR-T expansion and putative deleterious variants across the genome reveals significantly higher expansion among *PTPN22* variant carriers in both ZUMA-1 and ZUMA-7 a. Gene burden-based association testing with CAR-T peak expansion among all genes in ancestry-matched EUR patients. P-value by SKAT-O test. Results plotted by -log(p-value) for ZUMA-1 and ZUMA-7 with FDR and Bonferroni cutoffs as shown. b. CAR-T cell expansion metric comparison between *PTPN22* variant carriers and non-carriers. P-value by Mann-Whitney U-test. c. Heatmap correlating CAR-T cell expansion metrics between in silico pathogenicity predictors. CAR-T metrics were log transformed and normalized to 1 based on the maximum expansion within the cohorts. In silico pathogenicity predictors were normalized to 1 where appropriate.

## DISCUSSION

CAR-T cell therapy is a critical tool for the treatment of advanced hematologic malignancies, but inherited genetic drivers of their activity are still largely unexplored, representing an unmet need to better understand the underlying biological processes. In this study, we leveraged an integrative computational and functional approach to identify enrichment of putative deleterious variants among patients who had aggressive large cell lymphoma patients and had received axicabtagene ciloleucel in the ZUMA-1 and ZUMA-7 trials. Interrogating the complex phenotype of CAR-T cell-related toxicity and limiting our search to HLH-associated genes, we found enrichment of subjects with putative deleterious variants in *STXBP2* in ZUMA-1, and in vitro studies leveraging CRISPR-Cas9 to generate near-complete loss of function mutations, or the overexpression of variant STXBP2 proteins, were consistent with reduced degranulation and increased cytokine production. However, we did not find enrichment of *STXBP2* variants in subjects enrolled in the ZUMA-7 trial who experienced toxicity.

Broadening our search to the entire genome but this time including germline sequencing data from both clinical trials of axicabtagene ciloleucel, the most nominally significant finding across both cohorts with respect to toxicity was the enrichment of *ADAMTSL3* putative deleterious variants among control cohort subjects (i.e., those who did not experience severe toxicity), suggesting a possible protective effect from CAR-T associated toxicities. Finally, we found strong cross-cohort enrichment of *PTPN22* putative deleterious variant carriers among subjects with the highest levels of CAR-T expansion.

*STXBP2* plays a critical role in cytotoxic granule membrane fusion in lymphocyte granule-mediated cytotoxicity. Recently, a more complex picture of adult secondary HLH has emerged in which partial loss of function–hypomorphic or dominant negative (including p.A433V identified in this cohort)–variants in degranulation pathway members result in HLH-like manifestations with a sufficient immunologic challenge^26,28–33,37,47^. Germline variants in immune-related genes have also been associated with patients experiencing severe, hyperinflammatory manifestations of COVID-19^63^, including in *STXBP2^30^* and other perforin degranulation pathway members^64, 65^. Importantly, despite being referred to as ‘benign’ or ‘likely benign’ in clinical variant databases such as ClinVar, *STXBP2* putative deleterious variants from 4 of the 6 patients in our study have been recurrently identified in HLH patients^37,39–41,43,44^ and demonstrated to functionally impair degranulation^31,38,42^. We hypothesize that the engineered nature of CAR-T cell therapy, where high-affinity binders drive supraphysiologic T cell activation against many target cells, represents an extreme immunologic challenge for T cells and the immune system overall. This “stress test” may have the capacity to unmask or exacerbate hypomorphic protein functions that would otherwise have minimal or undetectable clinical penetrance.

Importantly, most of the toxicity in our combined toxicity endpoint was driven by high-grade neurologic toxicity rather than CRS, the former of which was more abundant in ZUMA-1 and ZUMA-7 overall. The toxicity in these subjects is similar to real-world outcome studies of individuals receiving axicabtagene ciloleucel outside the confines of a clinical trial^66, 67^. Overall, axicabtagene ciloleucel and its companion product, brexucabtagene autoleucel, are associated with higher rates of toxicity than other FDA-approved CAR-T cell therapies^20,68–70^, though there are specific nuances related to patient selection, trial design, and disease-specific interactions. We hypothesize that CAR-T CRS and HLH exist within a spectrum of immune and myeloid system activation. HLH-associated variants have been found to be associated with other hyperinflammatory contexts (e.g., Covid19^30, 71^) without necessarily resulting in overt HLH. Finally, current models of CAR-T cell neurotoxicity link central myeloid cell (microglia) activation as the key driver of this pathology^72, 73^. Degranulation defects in CAR-T cells could drive excessive myeloid activation systemically and in the central nervous system as a putative mechanism. The contributions of defects in HLH-associated genes in myeloid and other cell types remain unexplored in this study and may be important contributors to the pathophysiology of CRS and neurologic toxicity.

While *STXBP2* putative deleterious variants were enriched in the toxicity cohort in ZUMA-1, this was not seen in ZUMA-7. Importantly, ZUMA-1 was a much more heavily pre-treated group of patients, with 69% having received ≥3 lines of therapy and 77% having undergone an autologous stem cell transplant, versus ZUMA-7 patients, who were primary refractory or relapsed disease within 12 months and thus received only a single line of therapy. ZUMA-1 subjects were also more likely to have high-stage disease (85% vs 77% with stage III or IV disease). ZUMA-1 patients also had substantially higher levels of peak and AUC CAR-T cells in the toxicity cohorts than ZUMA-7 and higher baseline levels of inflammation. Given the rapidly evolving nature of toxicity management and grading, these findings do not invalidate our findings with respect to *STXBP2,* and additional larger studies will be needed to explore this complex association more fully, given the relative rarity of the discovery variants.

Leveraging the granularity of our correlative data, we found strong associations between CAR-T cell expansion, most notably with *PTPN2*2 variant carriers who had the highest expansion in both of our cohorts, despite making up just 5% of all subjects. While disruption of *PTPN22* in T-cells and across the entire organism has been demonstrated to augment anti-tumor responses^11^, the impact of our other identified genes remains to be explored.

One limitation of our findings is that the potential generalizability to other CAR-T cell designs, target antigens, tumor types, or ancestral populations will need to be confirmed in future studies. Furthermore, we used CRISPR-Cas9 to generate near-complete loss-of-function mutations or the overexpression of variant proteins, which may not exactly recapitulate the behavior of the individual heterozygous variants we identified in patients.

Unlike most traditional pharmacologic compounds in which there is a single, molecularly defined “active ingredient,” autologous CAR-T cells are bespoke, living drugs that carry all the ancestral polymorphisms of the individual from whom they are manufactured. This has important implications for CAR-T cell behavior. Allogeneic CAR-T (allo-CAR-T) cells, in which a single healthy donor’s T cells may be used to manufacture CAR-T cells for potentially several individuals, are currently being tested in numerous clinical trials^74^. Our work suggests that germline-level donor screening may have implications for product behavior since a single donor may provide T cells sufficient for dozens to hundreds of CAR-T cell products.

In summary, our study establishes the first clear association between germline variants and CAR-T cell behavior. We anticipate that this highly annotated dataset, incorporating whole genome sequencing matched to two clinical trials of CAR-T-treated aggressive lymphoma patients, will provide further valuable insights into the relationship between germline variants and engineered immune cell behavior.

## Data Availability

The data that support the findings of this study are available from Kite Pharma, but restrictions apply to the availability of these data, which were used under license for the current study, and so are not publicly available due to patient privacy concerns. Data are however available from the authors upon reasonable request and with permission of Kite Pharma.

## METHODS

### Study design and population of patients

We obtained available genomic DNA from patients in the ZUMA-1 and ZUMA-7 clinical trials. All patients had provided written informed consent as part of the ZUMA-1 and ZUMA-7 studies at their respective institutions. None of the authors of this manuscript had access to identifiable patient information and nor did they participate in the consent process for the individuals. We stratified the patients *a priori* into a case-control study design, with the toxicity (case) cohort defined as individuals who had severe (Grade 3 or higher out of 5, with higher scores indicating more severe toxicity) cytokine release syndrome (CRS; graded by Lee criteria^75^) or neurotoxicity (graded by CTCAE 4.03) or had received the interleukin-6 (IL-6) cytokine receptor blocker tocilizumab with the presumption that the latter represented a proxy for clinically significant CRS. The remainder (Grade 0-2 CRS or neurotoxicity and did not receive tocilizumab) were designated the control cohort. No patients had previously undergone allogeneic stem cell transplant.

### Whole genome sequencing and Evaluation of Sequencing Read Coverage

Genomic DNA (gDNA) was extracted from peripheral blood mononuclear cells (PBMC) of all de-identified subjects, and we performed whole genome sequencing at the Broad Institute (Cambridge, MA) to a mean sequencing depth of at least 30X on an Illumina HiSeqX (Illumina, San Diego, CA) with 150 base paired-end reads. Samples with sufficient total DNA quantity underwent PCR-free WGS (ZUMA-1: n=84, ZUMA-7: n=134) while samples with relatively low DNA quantity underwent PCR amplification before WGS (ZUMA-1: n=2, ZUMA-7: n=16). Raw sequencing Binary Alignment Map (BAM) files were aligned to the human reference genome GRCh38 using BWA (version 0.7.15).^76^ The average sample-level sequencing coverage for exome-wide intervals was calculated using GATK^77^ (version 3.7) tool “DepthofCoverage” to ensure sufficient read depth to make confident germline calls. BAM files for all samples included in the analysis had a sample mean coverage of over 25X across the genome.

### Germline Sequencing metrics

All samples obtained mean coverage in HLH-associated genes greater than 25x and performance characteristics were similar between the cohorts. Germline variants were identified using DeepVariant and pathogenicity was classified according to AlphaMissense^78^, Combined Annotation Dependent Depletion (CADD v1.7)^79^, Mendelian Clinically Applicable Pathogenicity (M-CAP) ^80^, MetaLR^81^, MetaRNN^82^, Polymorphism Phenotyping, version 2 (Polyphen-2)^83^, PrimateAI^84^, Rare Exome Variant Ensemble Learner (REVEL)^85^, Sorting Intolerant from Tolerant (SIFT) ^86^ We selected a minor allele frequency of less than 0.05 across populations to define rare variants.

### Ancestry Inference

Case and control cohort-level VCF files were combined with the cohort-level VCF of 1,000 Genomes project reference samples^87^ (n=2,504) with known continental ancestries of Admixed American (AMR), African (AFR), European (EUR), East Asian (EAS), and South Asian (SAS). The combined file was filtered for germline variants in the highly covered exome intervals (>15x). Then the filtered VCF file was loaded into a “Hail” matrix table (version 0.2.11, https://github.com/hail-is/hail), and rare germline variants of a cohort-level allele frequency below 1% and deviating from Hardy-Weinberg equilibrium (chi-squared p-value <1×10^-6^) were excluded. Then, we performed linkage disequilibrium (LD) pruning using the “ld_prune” method in Hail to exclude variants with Spearman correlation coefficient >0.1 within a 1 million base pair window. These filtered germline variants were used for PCA with the Hail “hwe_normalized_pca” method. Sklearn package (version 0.20.0) “RandomForestClassifier” function was then applied to the top 10 global principal components of reference samples from 1000 Genomes Project to train random forest classifiers for the five continental ancestries. This was used to assign broad continental ancestry to the cases and controls. Given the limited number of non-EUR patients, only EUR samples were included for all downstream analyses.

### Methods of germline variant calling, relatedness analysis, and prioritization of variants are detailed in the Supplementary Information

#### Functional and Clinical Annotation and Prioritization of Germline Variants

A curated list of HLH-associated genes previously interrogated in an adult population (**Extended Data Table S1**)^6^ was used to identify candidate variants. All identified variants were then classified into five categories: benign, likely benign, variants of unknown significance, likely pathogenic, and pathogenic, according to the American College of Medical Genetics (ACMG) guidelines using ACMG Classification.^88^ For our primary case-control analysis we focused on “putative deleterious” variants, which include variants that have strong evidence for loss of function (i.e. frameshift, stop codon, and canonical splice variants), those that are annotated as pathogenic or likely pathogenic in ClinVar, and rare missense variants with minor allele frequency (MAF) (<0.05) in The Genome Aggregation Database (gnomAD). The presence of the identified germline variants in *STXBP2* was further validated by manual examination of the BAM files using Integrative Genomics Viewer (IGV, version 2.11.1).^89^ Separately, we investigated the enrichment of rare (gnomAD max AF <5%) synonymous and common (gnomAD max allele frequency>=5%) missense variants as control analyses. For the purposes of gene-level burden testing, patients with at least one variant in the gene were considered to be ‘positive’ and were compared to those with no variants in a given gene.

## Statistical Analysis and Data Visualization

Exact2×2 R package was used to calculate odds ratios, 95% confidence intervals, and p-values for two-sided Fisher’s exact test for gene burden analysis. Q-values for multiple hypothesis testing were computed using the R package “qvalue” (http://github.com/jdstorey/qvalue). Binomial proportional confidence intervals were calculated using the following web-based tool (https://sample-size.net/confidence-interval-proportion/). The R package “SKAT” (version 2.2.4 https://cran.r-project.org/web/packages/SKAT/index.html) was used for SNP-set (Sequence) Kernel Association Test (SKAT) testing. Burden-based testing figures were generated using RStudio (Version 2022.07.2) via Jupyter notebook (version 6.4.6) and Python “Matplotlib” package (version 3.10.1) ^90^. R “ggplot” library was used to generate figures to visualize variant data. The “co-mutation” plot summarizing the demographic and clinical information as well as the variant status of the cohort was generated using Python “CoMut” package (version 0.0.3, https://github.com/vanallenlab/comut).^91^ *In vitro* experiment statistics were calculated using “Prism” version (9.4.1) with the indicated statistical test in the figure legend. Adobe Illustrator (version 26.0.2) was used for final figure preparation.

The methods for CAR design, CAR-T cell production for *in vitro* testing and validation, as well as functional assays, are described in the Supplementary Appendix.

## Acknowledgements

This study was funded by Kite Pharma. Nalm6-BID KO cells were a gift from Dr. Nathan Singh.

## Author Contributions

MBL, MVM, SA, EVA, RRS, and KMG designed the study. MBL, KMG, HS, AB and GM performed the experiments. MBL, BS, SH, JB, and HC analyzed the data. MBL compiled the manuscript, which was edited by MVM, SA, BS, RRS, and SF. All authors contributed intellectually to the project and reviewed the manuscript and approve of its content.

## Competing interests

JB, SF, and RRS are Kite Pharma employees. M.V.M is an inventor on patents related to adoptive cell therapies, held by Massachusetts General Hospital and the University of Pennsylvania (some licensed to Novartis). Dr. Maus holds equity in TCR2, Century Therapeutics, Genocea, Oncternal, and Neximmune, serves on the Board of Directors of 2Seventy Bio, and has served as a consultant for multiple companies involved in cell therapies. MVM’s interests were reviewed and are managed by Massachusetts General Hospital, and Mass General Brigham in accordance with their conflict-of-interest policies. BDC, MVM, and MBL are inventors of patents related to the use of engineered cell therapies. BDC received commercial research grants from ACEA Biosciences. R.G. has equity in Moderna, Pfizer, and Vertex Pharmaceuticals.

Supplementary Information is available for this paper.

---

**Extended Data Figure 1.**
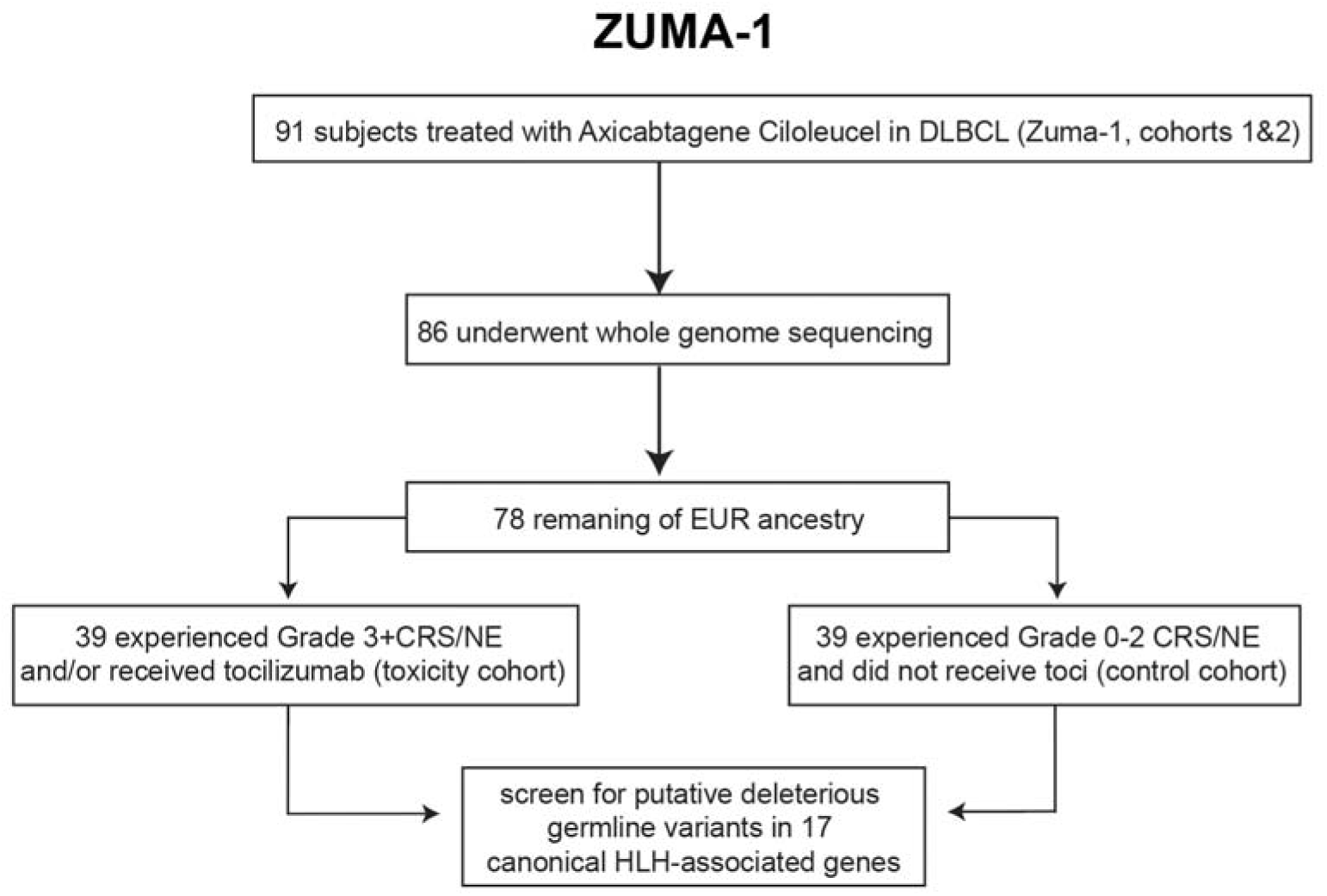
Study design Germline analysis cohort availability and workflow. Only subjects from cohorts 1 and 2 were included (criteria for tocilizumab administration changed for later cohorts and thus were excluded).

**Extended Data Figure 2.**
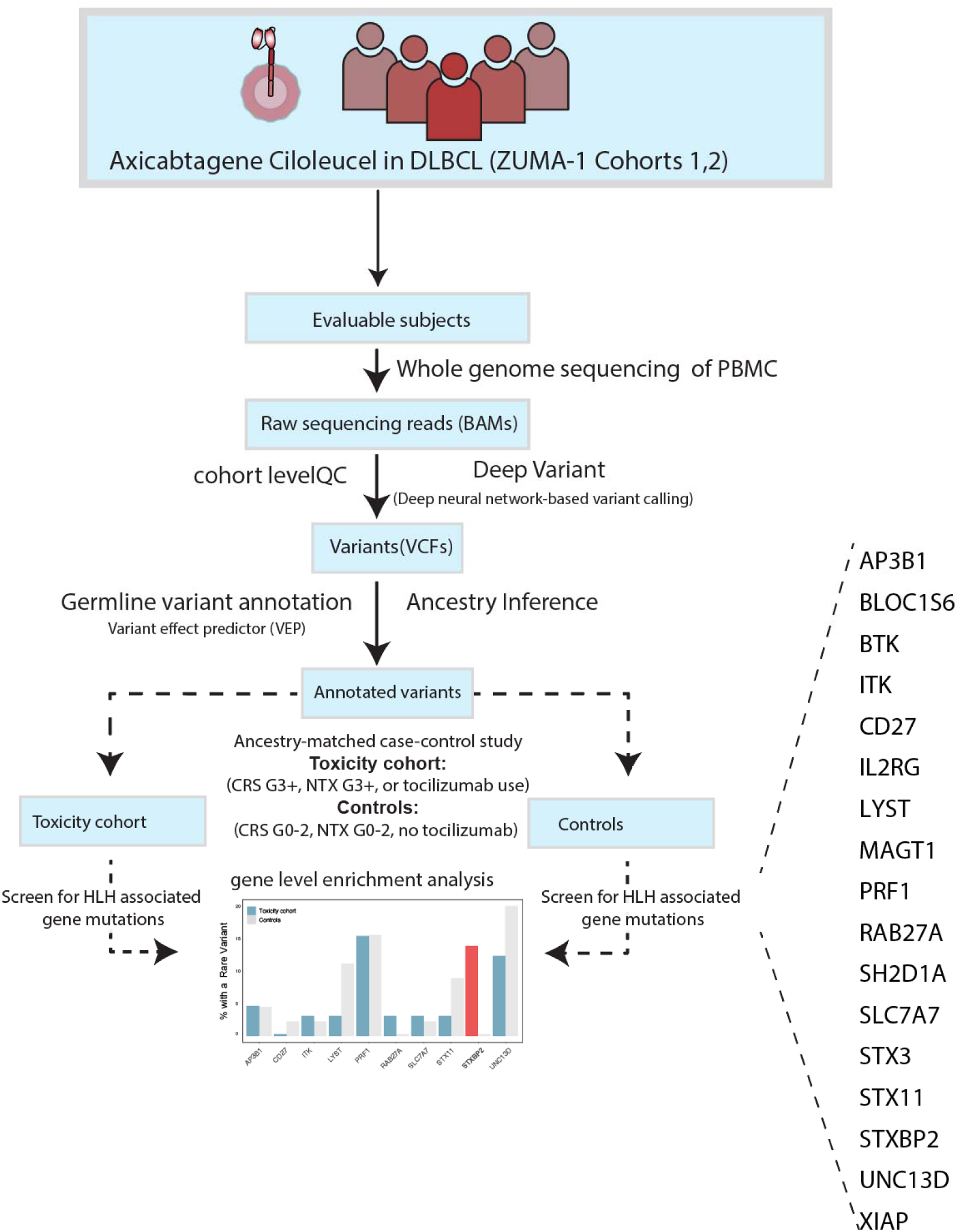
Detailed study outline Included patients, analysis steps, and workflow are shown

**Extended Data Figure 3.**
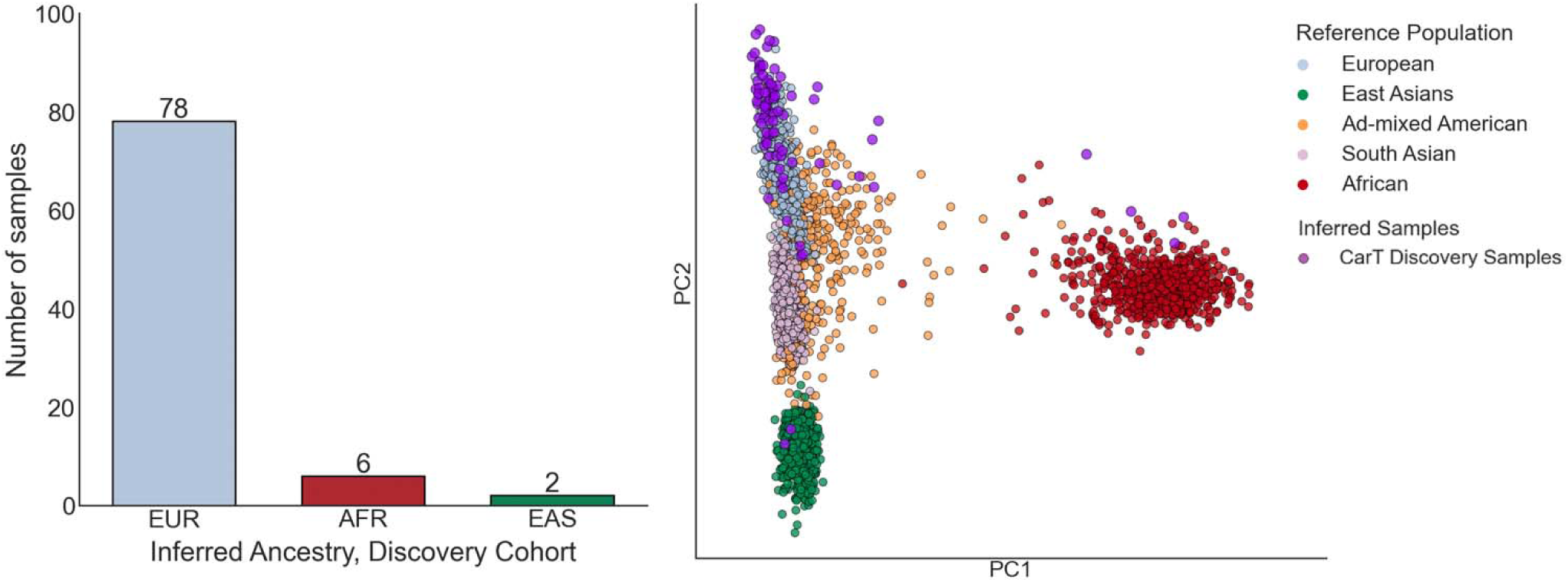
Ancestry inference Principal component analysis (PCA)-based ancestry inference results (right) and continental ancestry assignments (left).

**Extended Data Figure 4.**
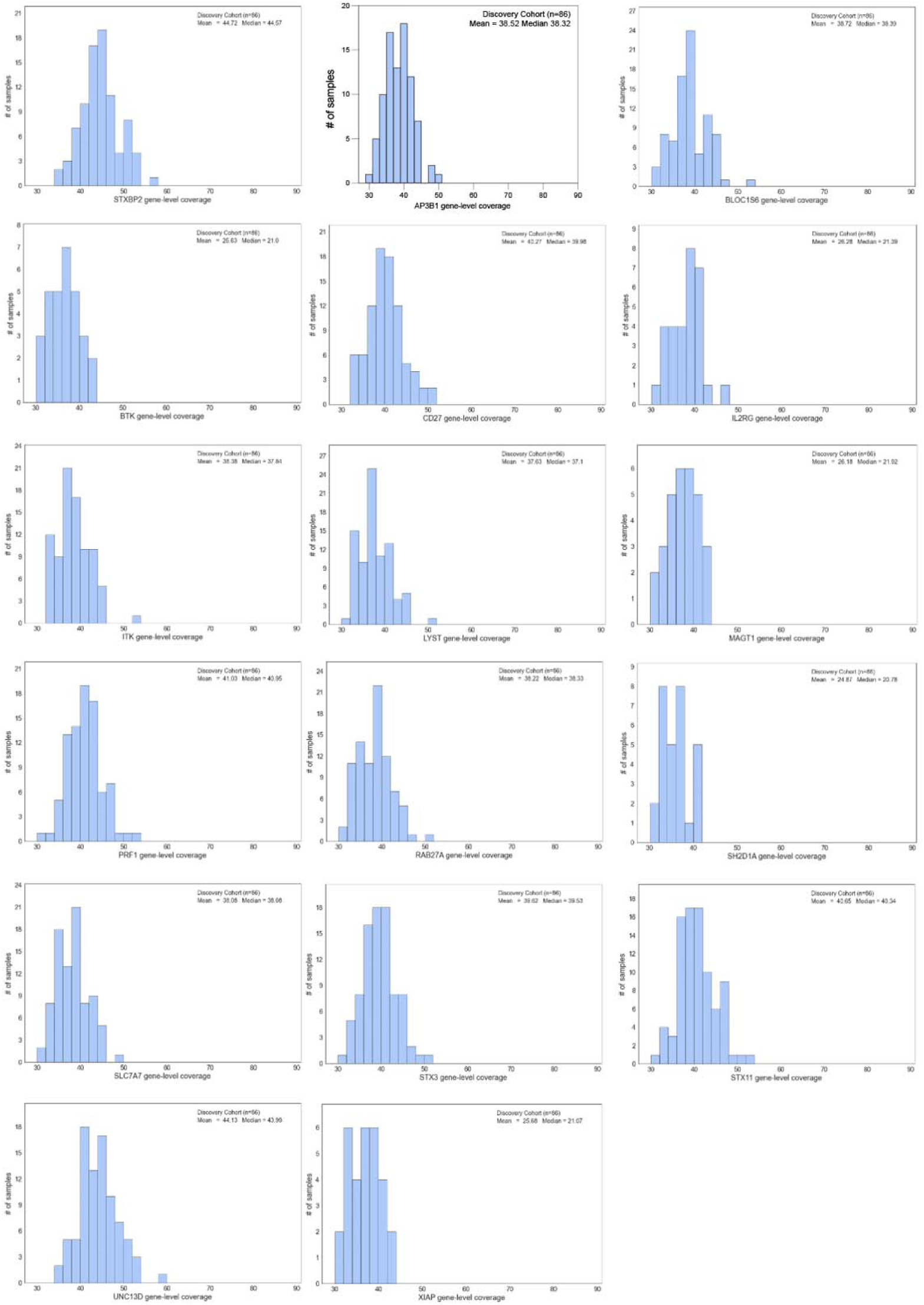
HLH gene coverage depth Coverage statistics for the 17 genes included in this study

**Extended Data Figure 5.**
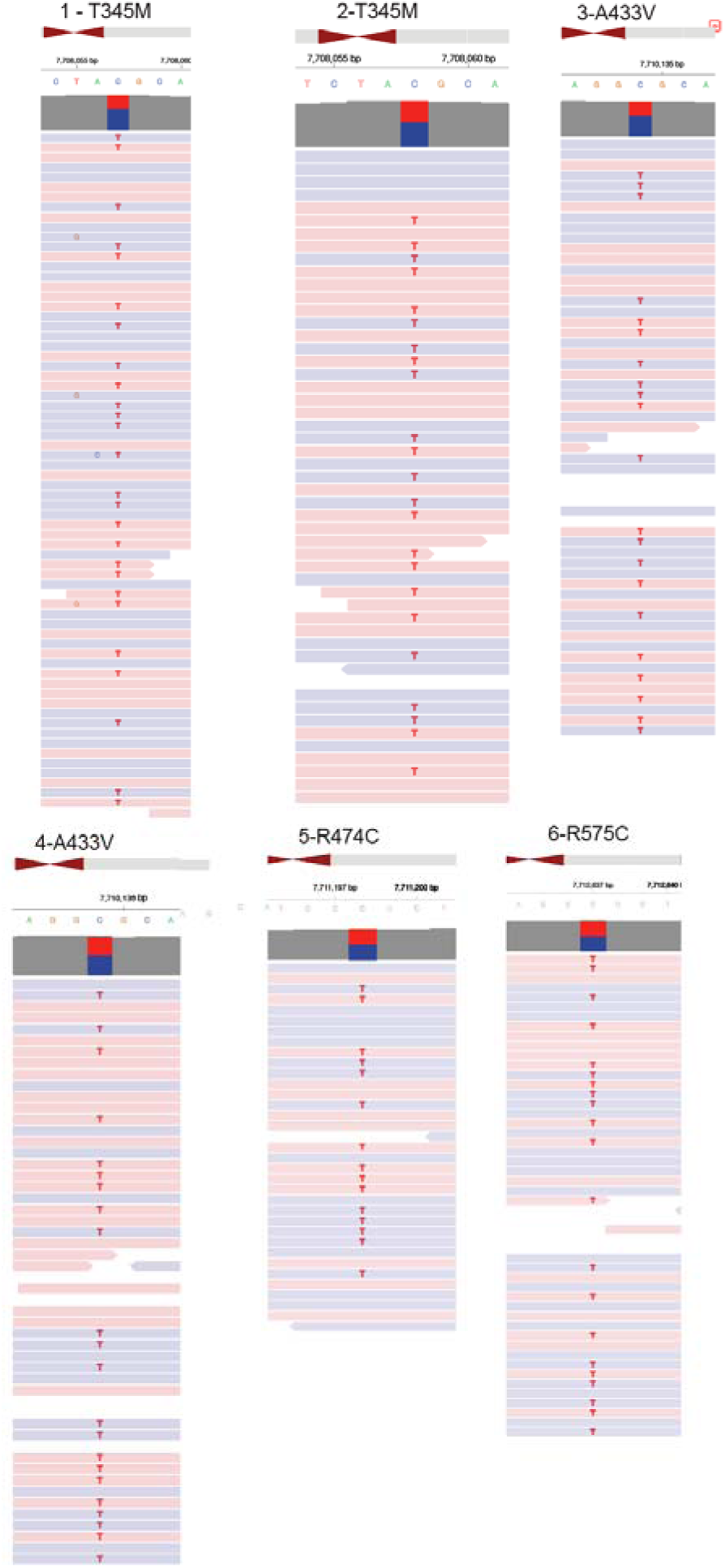
Integrated Genomics Viewer (IGV) snapshots of putative deleterious *STXBP2* variants. Patient ID and *STXBP2* putative deleterious variants are identified. Colors represent paired read strand direction.

**Extended Data Figure 6.**
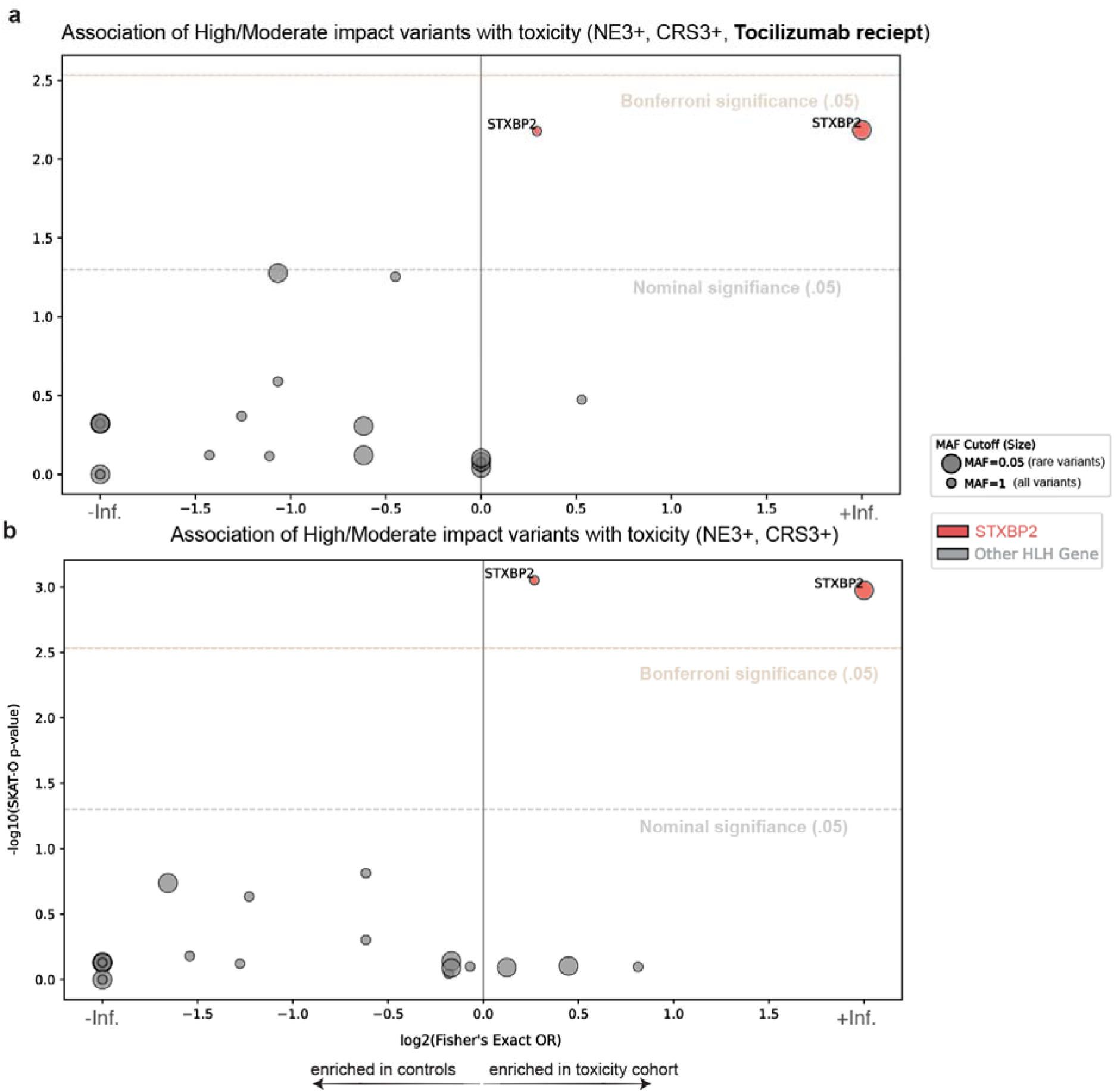
Effect of the receipt of tocilizumab on the combined toxicity endpoint. Putative deleterious variants were identified with the indicated MAF thresholds and plotted against the SKAT-O p-value. a. Analysis using high-grade CRS (3+), neurotoxicity (NE, 3+), or receipt of tocilizumab to establish the toxicity cohort b. Analysis using only high-grade CRS (3+), or neurotoxicity (NE, 3+) to establish the toxicity cohort. Rare variant analysis results (MAF < 0.05) with tocilizumab: p=6.52e-3, FDR=0.143, Bonferroni-adjusted p=0.111; excluding tocilizumab: p=1.06e-3, FDR=0.0233, Bonferroni-adjusted p=0.0180).

**Extended Data Figure 7.**
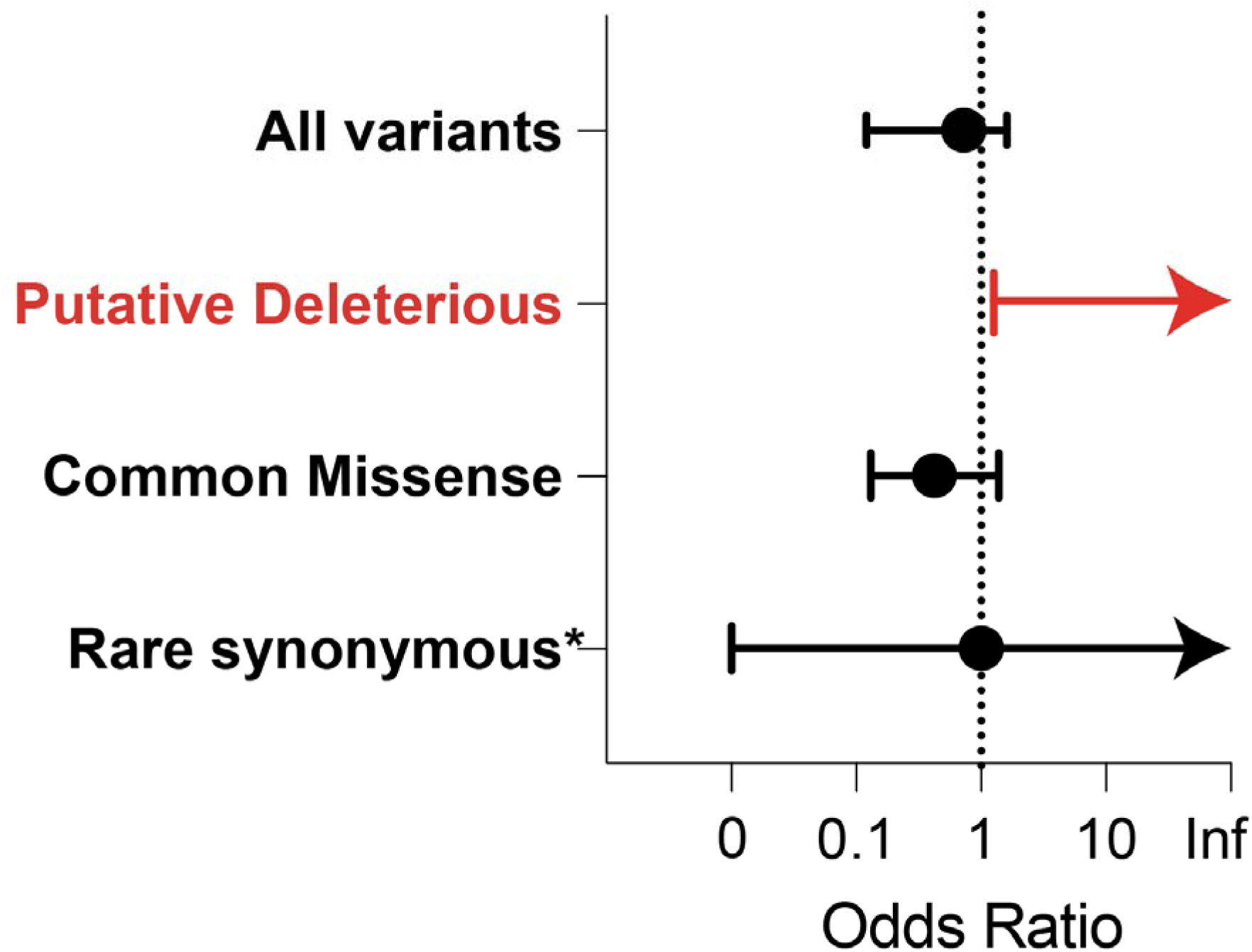
Comparison of *STXBP2* odds ratio association with toxicity endpoint and variant types Odds ratio with 95% confidence interval shown. *Only a single rare synonymous variant was identified.

**Extended Data Figure 8.**
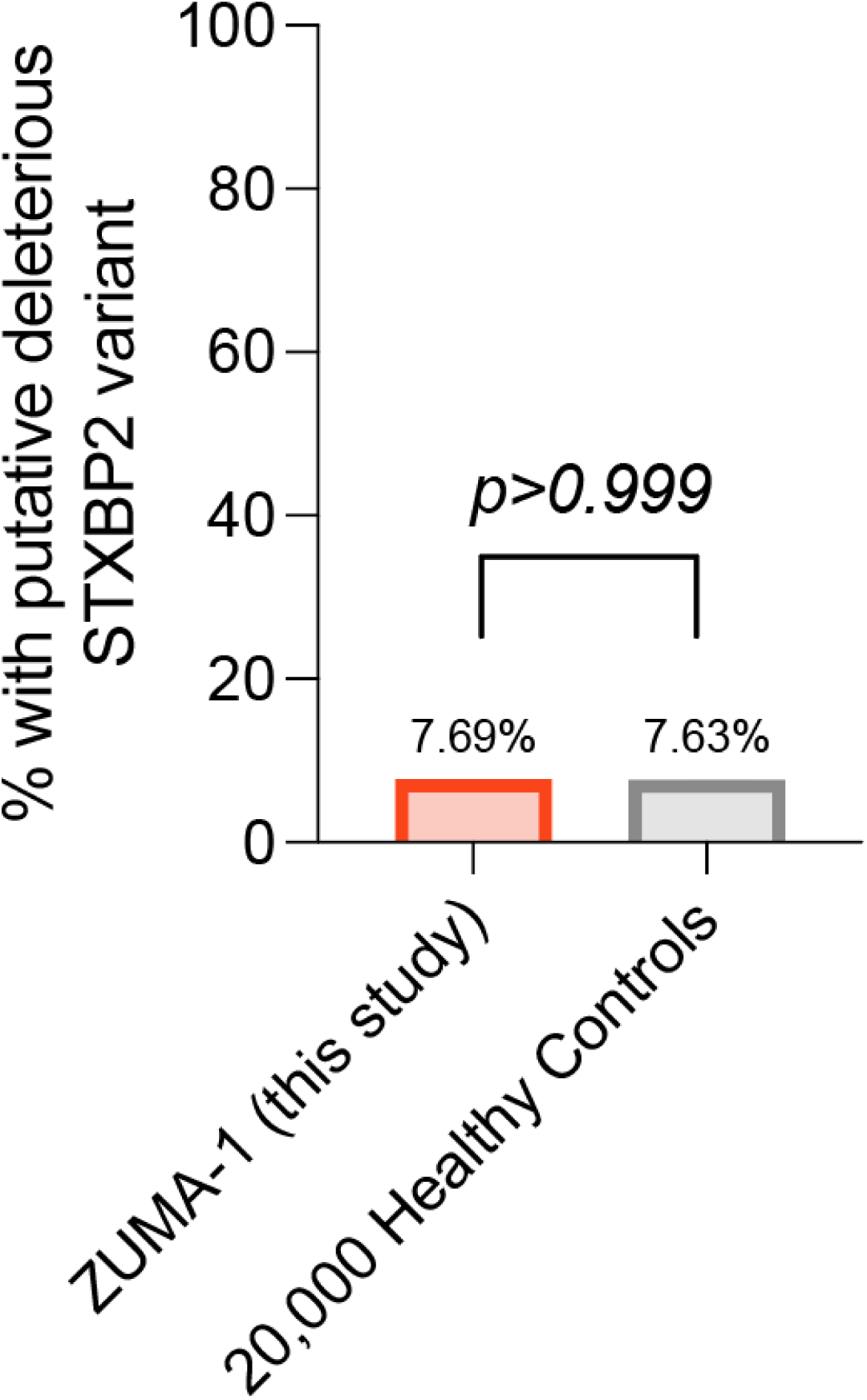
Prevalence of *STXBP2* putative deleterious variants in healthy control subjects. Percentage of subjects with an *STXBP2* putative deleterious variant in this study compared to an ancestry matched cohort of individuals without a cancer diagnosis. P-value represents a two-tailed fisher’ exact test.

**Extended Data Figure 9.**
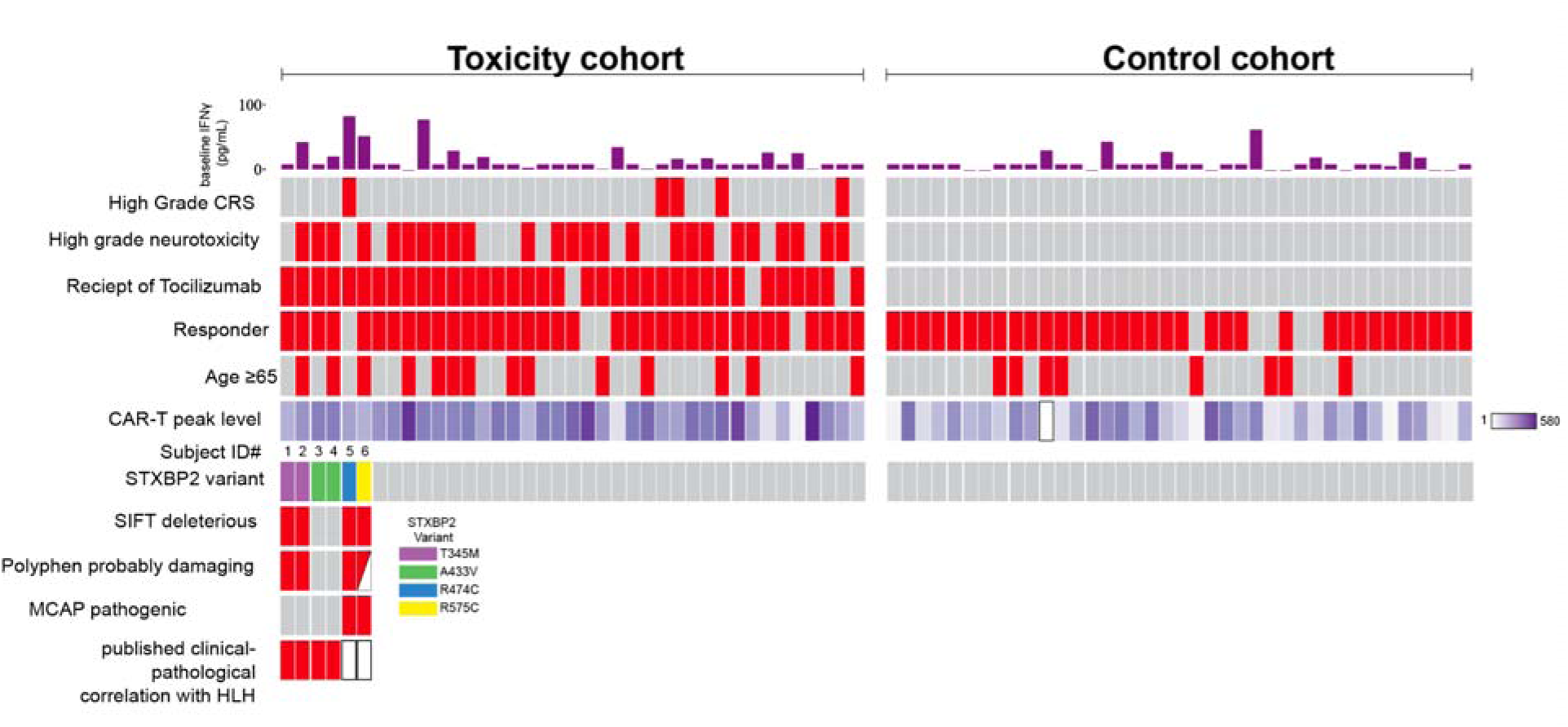
Co-mutation plot showing association between demographic, clinical, and variant characteristics Each column represents an individual patient in the cohort. Patients are stratified by cohort. Baseline IFNγ is plotted (top row). Features of the combined toxicity endpoint (Grade 3 or higher CRS or neurotoxicity (NTX), or receipt of tocilizumab) are annotated. Best response and age (less than or greater than or equal to 65) are also shown. CAR-T peak expansion (cells/µL) is shown as a heatmap. Specific *STXBP2* variants, along with *in silico* pathogenicity predictions, are shown along with annotations for prior published associations with pathogenicity. Half-filled boxes represent intermediate values of pathogenicity predictors. Demographic, variant, and clinical variables are displayed. CRS-cytokine release syndrome, NTX-neurotoxicity, SIFT-sorting intolerant from tolerant, Polyphen-Polymorphism Phenotyping v2, MCAP-mendelian clinically applicable pathogenicity

**Extended Data Figure 10.**
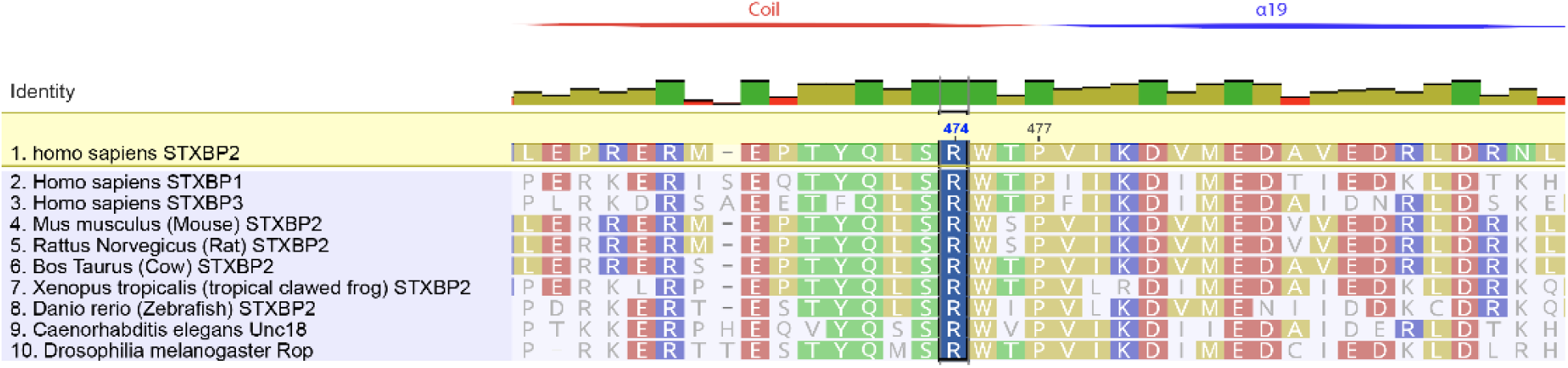
Conservation of R474 across *STXBP* family members and species. *STXBP2* R474 is highlighted, as well as adjacent P477, in which P477L was found to define FHL-5^45^. Different *STXBP2* family members and species homologs are aligned to human *STXBP2*.

**Extended Data Figure 11.**
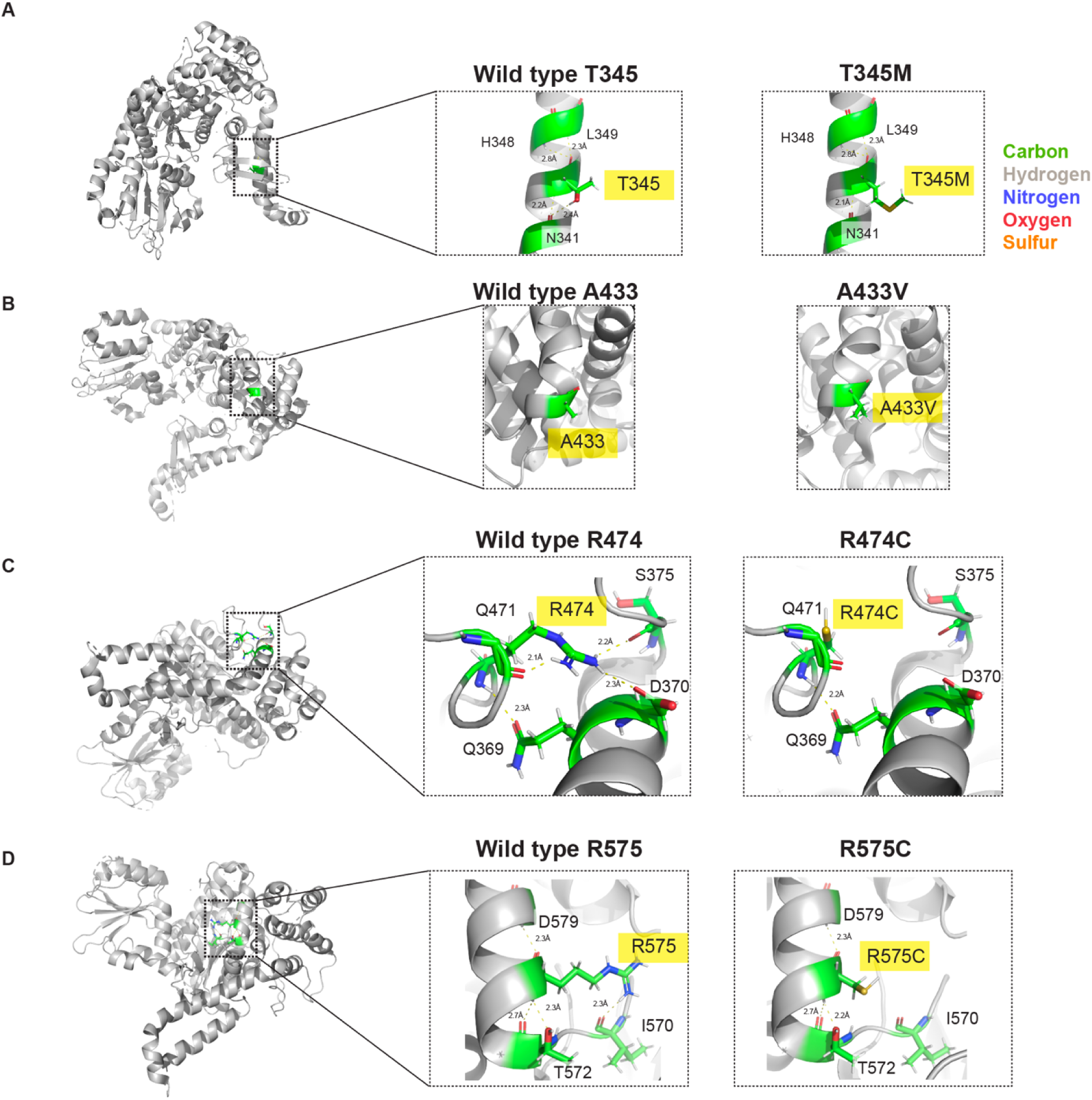
Protein modeling of *STXBP2* variants The sequence of *STXBP2* was obtained from the protein data bank (4cca). Pymol was utilized to visualize the variants identified in the study. Domains were imputed as in **Fig. S7** from *STX11*. **A**. T345M occurs within an alpha helix region of domain 3a and results in the loss of a hydrogen bond with N341 which was predicted to be the destabilizing and the cause of pathogenicity by Hackmann et al. during solving of the crystal structure.^41^ **B.** A433V occurs within domain 3b near the terminus of an alpha helix. Substitutions for valine are generally poorly tolerated within alpha helices, given valine’s limited ability to form the requisite bond angles.^92^ **C.** R474C occurs within domain 3b, resulting in the loss of salt bridge formation with nearby D370 on an alpha helix, as well as adjacent Q471 and S375. **D.** R575C occurs in domain 2 near the carboxy terminus of the protein. Variant results in the loss of a salt bridge formed between adjacent I570 on a loop structure.

**Extended Data Figure 12.**
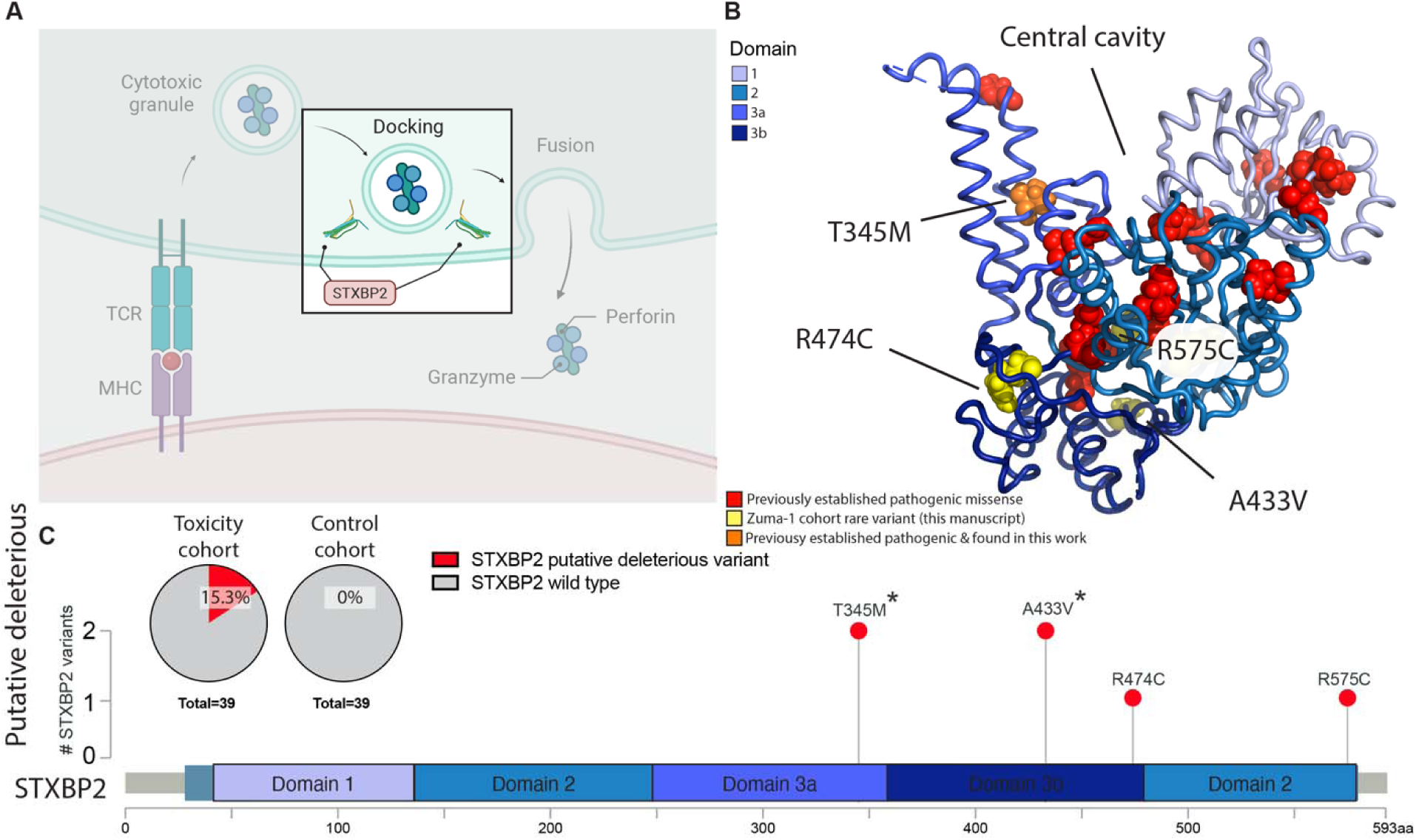
*STXBP2* function and pathogenic variant distribution **A**. T cell degranulation pathway with the role of *STXBP2* highlighted **B**. Crystal structure of *STXBP2* ^41^ overlaid with functional domains from the homolog *STXBP1^93^* and colored with variants modeled by Hackmann et al. (red), those found in this manuscript (yellow) and those found in both (orange) **C**. Putative deleterious variants of *STXBP2* mapped on the linear protein structure.

**Extended Data Figure 13.**
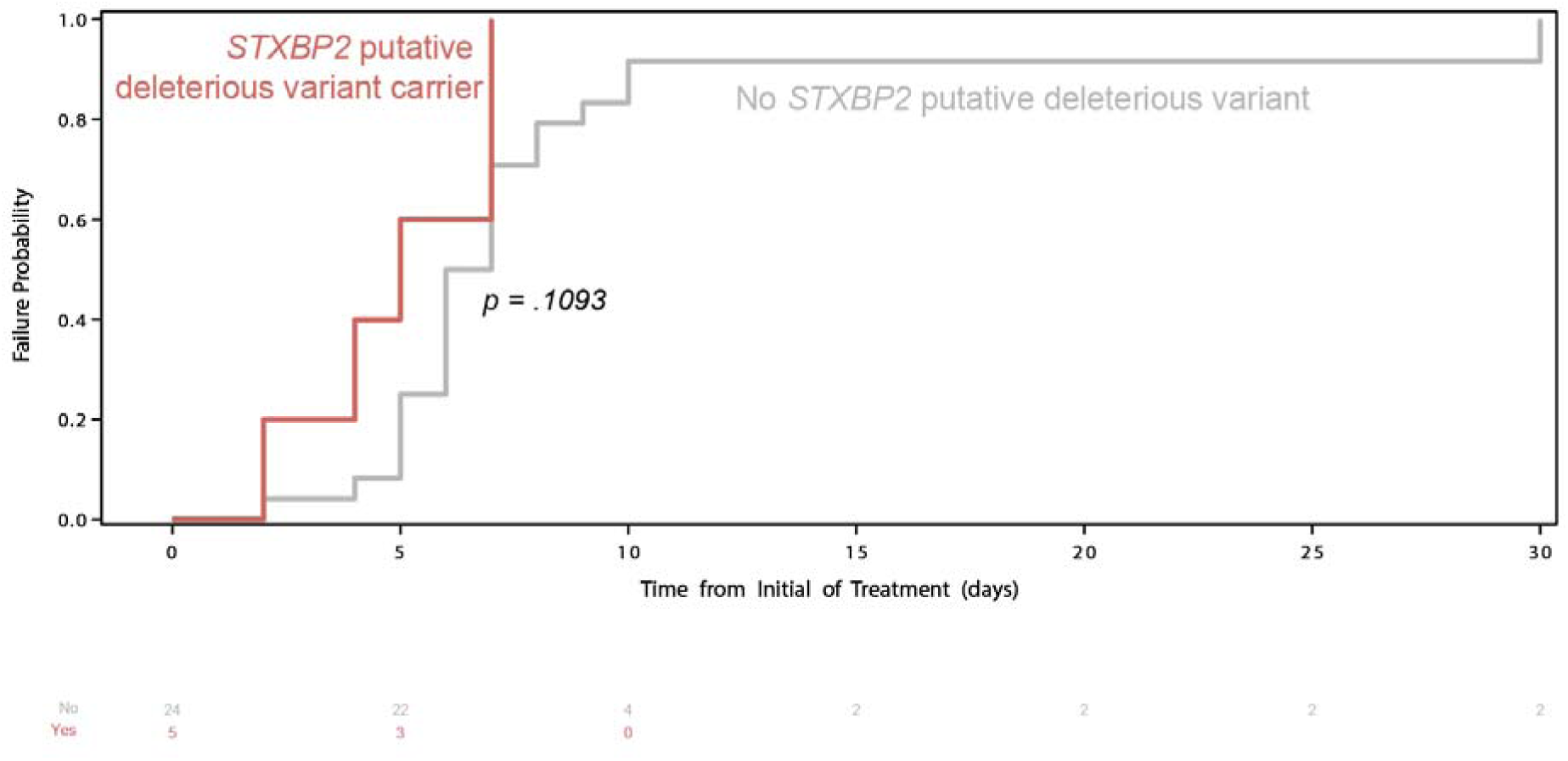
*STXBP2* variant carriers’ trend towards earlier onset of high-grade CRS and neurologic toxicity Event curve displaying attainment of high-grade CRS or neurotoxicity for STXBP2 putative deleterious variant carriers versus the remainder of the toxicity cohort. Patients only met the combined toxicity endpoint through the receipt of tocilizumab were excluded. P-value calculated by Log-rank (Mantel-Cox) test.

**Extended Data Figure 14.**
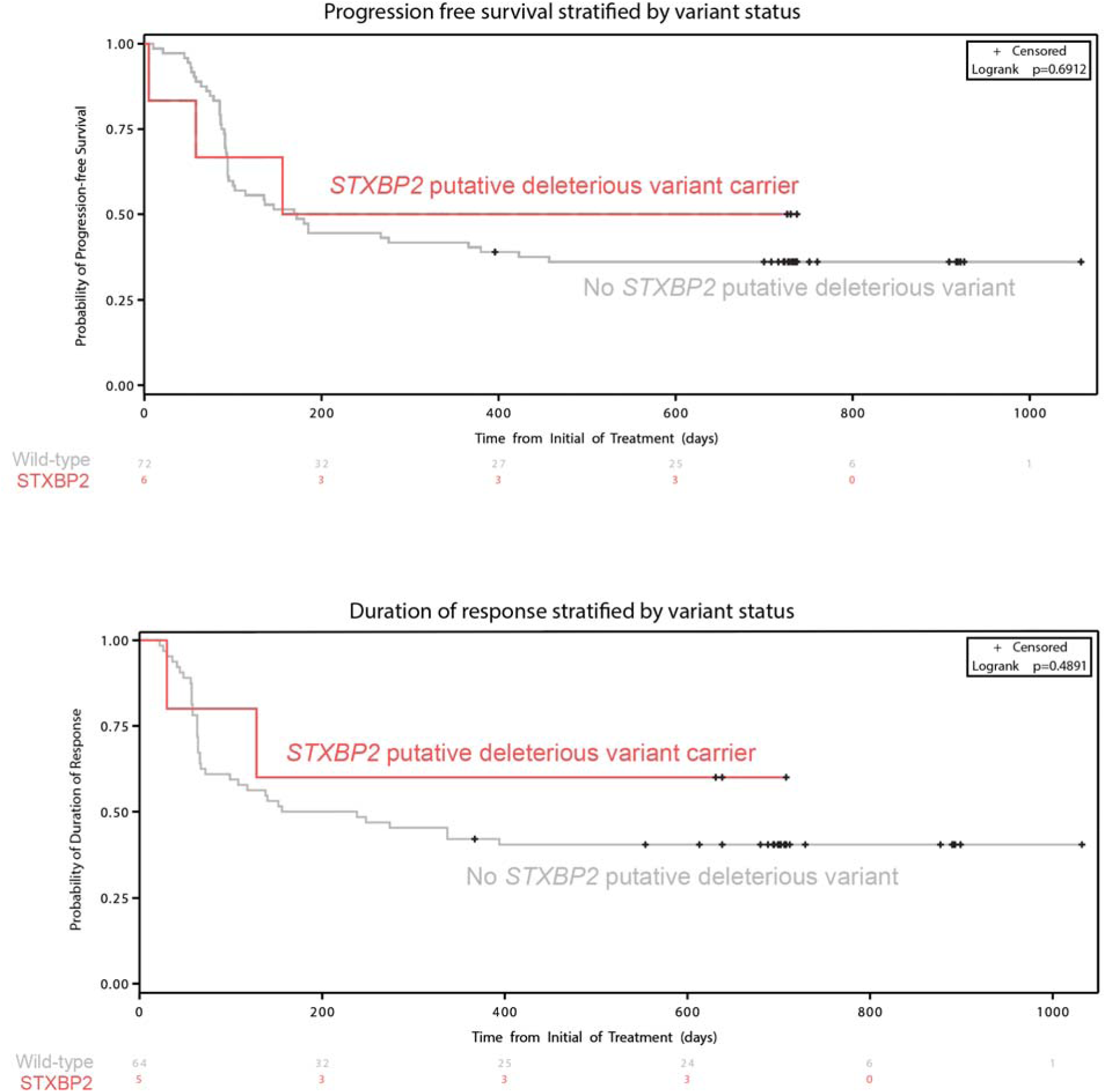
Progression free survival and duration of response of variant carriers. PFS (top) and DOR (bottom) of *STXBP2* putative deleterious variant carriers versus non-carriers from the entire study are compared. P-value represents logrank test.

**Extended Data Figure 15.**
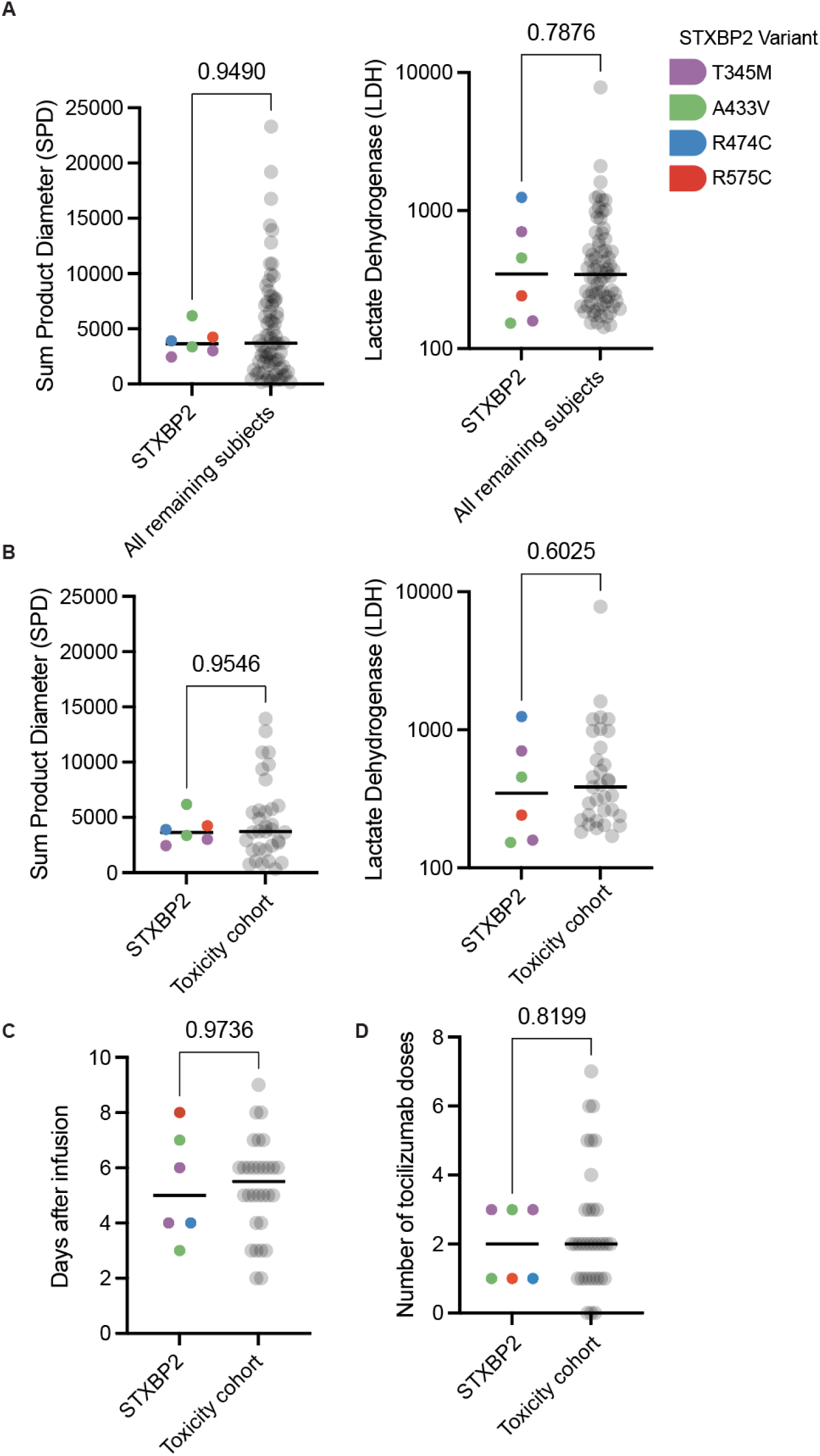
Baseline tumor and CRS treatment characteristics of *STXBP2* putative deleterious variant carriers. **A-B.** Baseline sum product diameter (SPD) measurement^94^ of tumor of *STXBP2* putative deleterious variant carriers or lactate dehydrogenase (LDH) versus all remaining subjects (**A)** or the remainder of the toxicity cohort **(B). C.** Day after CAR-T infusion of first tocilizumab administration of variant carriers versus remaining subjects who received tocilizumab**. D.** Total number of tocilizumab doses after CAR-T infusion. P-values represent individual Mann-Whitney U tests.

**Extended Data Figure 16.**
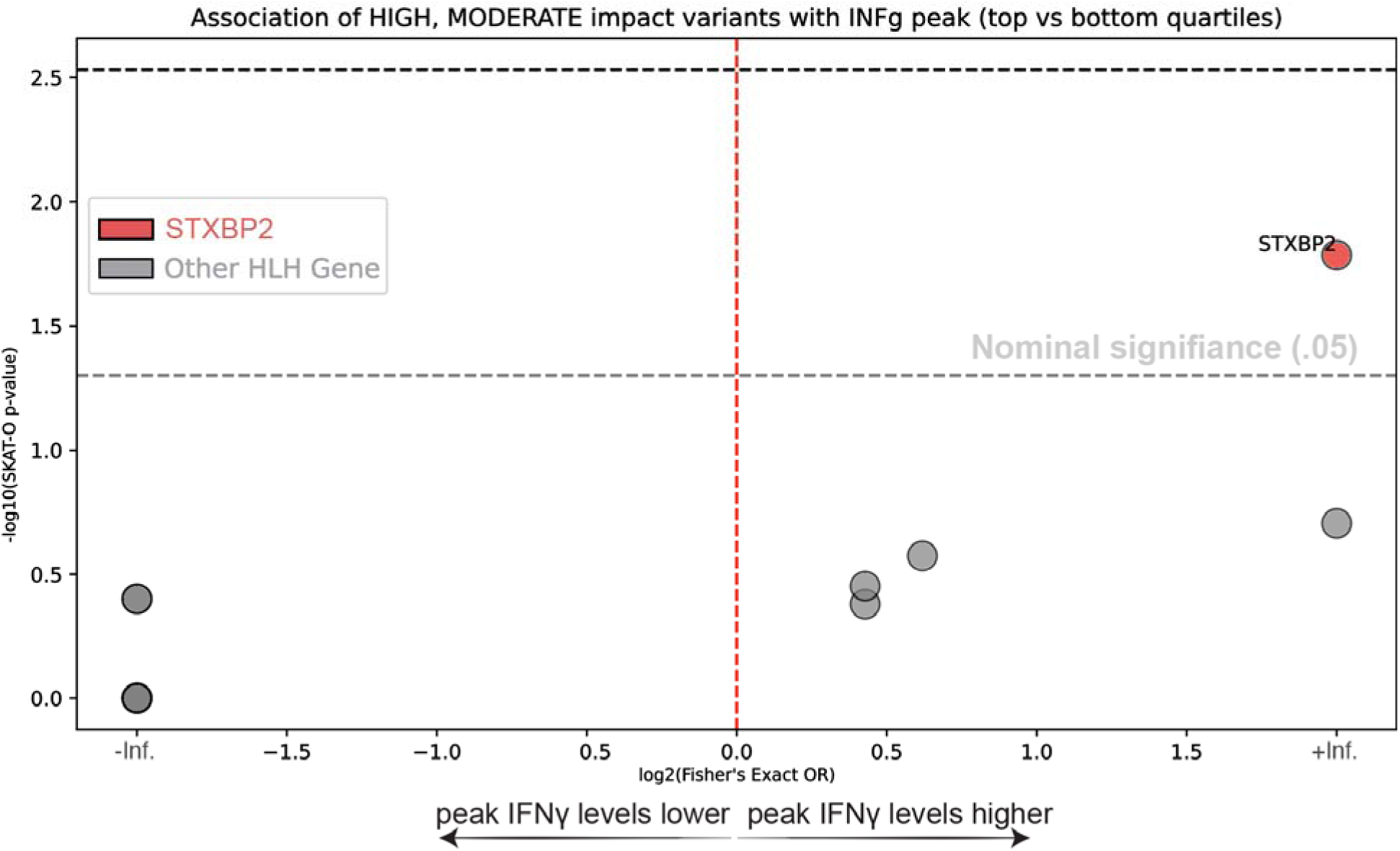
Association between HLH-associated genes and IFNγ levels. Analysis of putative deleterious variants in HLH-associated with peak IFNγ levels within patients with the highest and lowest IFNγ levels in the cohort, defined by top and bottom quartiles of peak IFNγ. STXBP2 was found to be associated with higher peak IFNγ levels (p-value = 0.0164). P-value by SKAT-O. inf-infinity

**Extended Data Figure 17.**
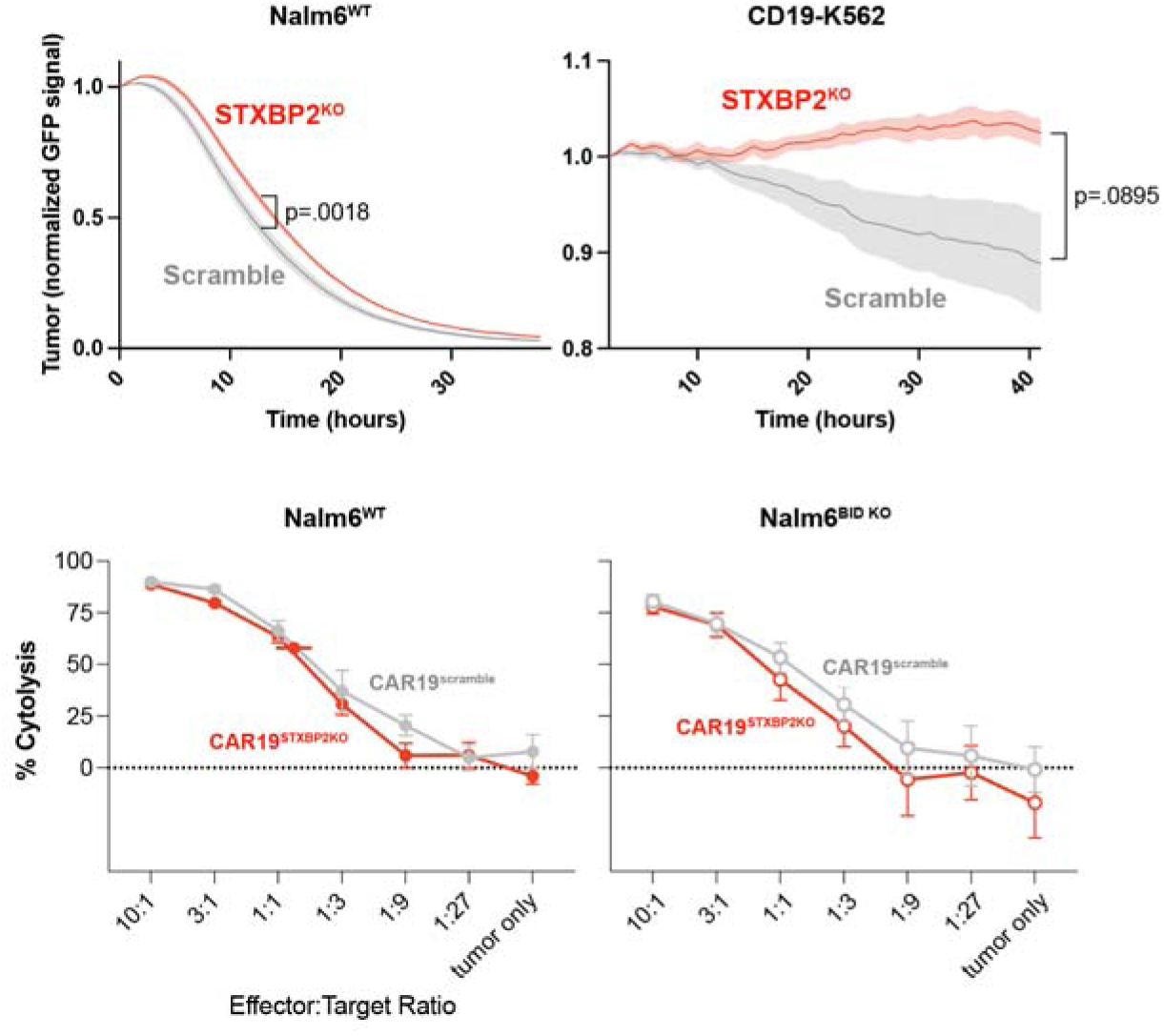
CD19-targeted *STXBP2*KO CAR-T cells have impaired *in vitro* cytotoxicity Real time cytotoxicity assay comparing *STXBP2* knockout and scrambled guide control CAR-T cells against indicated cell lines. Data points inclusive of three unique CRISPR guides and CAR-T cells from two donors. Mean± SEM is shown. Bottom, luciferase-based killing assay over 18 hours against the indicated targets and at the shown effector to target ratios. n=2 donors in technical triplicate. P-value represents 2way ANOVA.

**Extended Data Figure 18.**
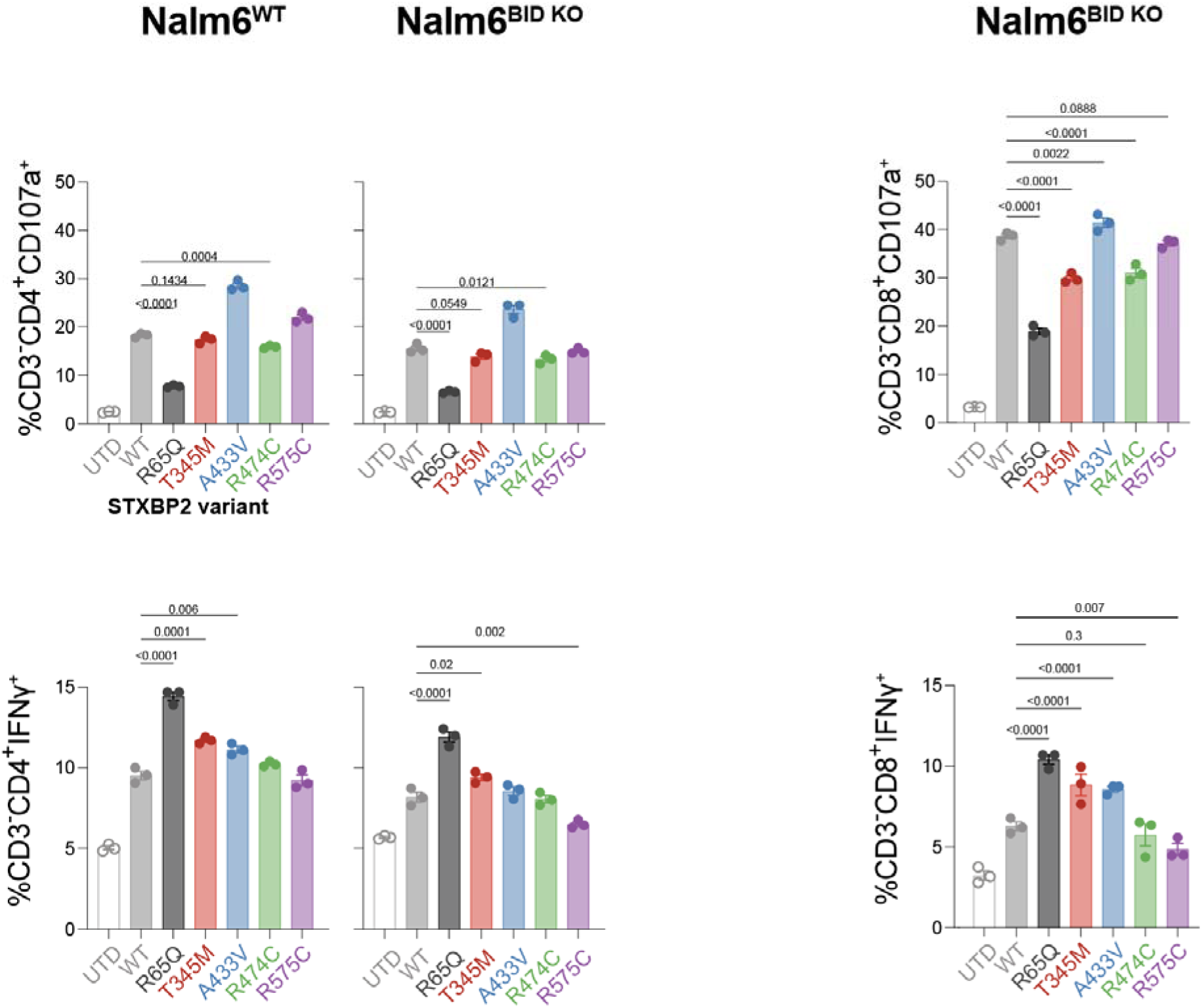
*STXBP2* putative deleterious variants impair degranulation and drive increased IFNg production Additional data related to **Fig. 2k,l**. Primary T-cells transduced with the constructs in 2k. were expanded and selected for edited T-cells prior to use in a 4-hour co-culture with Nalm6 or Nalm6-BID knockout leukemia target cells^95^. Cells then underwent flow cytometry for the indicated markers with representative flow plots. Bars represent the mean of technical triplicates. P-values represent 2way ANOVA.

**Extended Data Figure 19.**
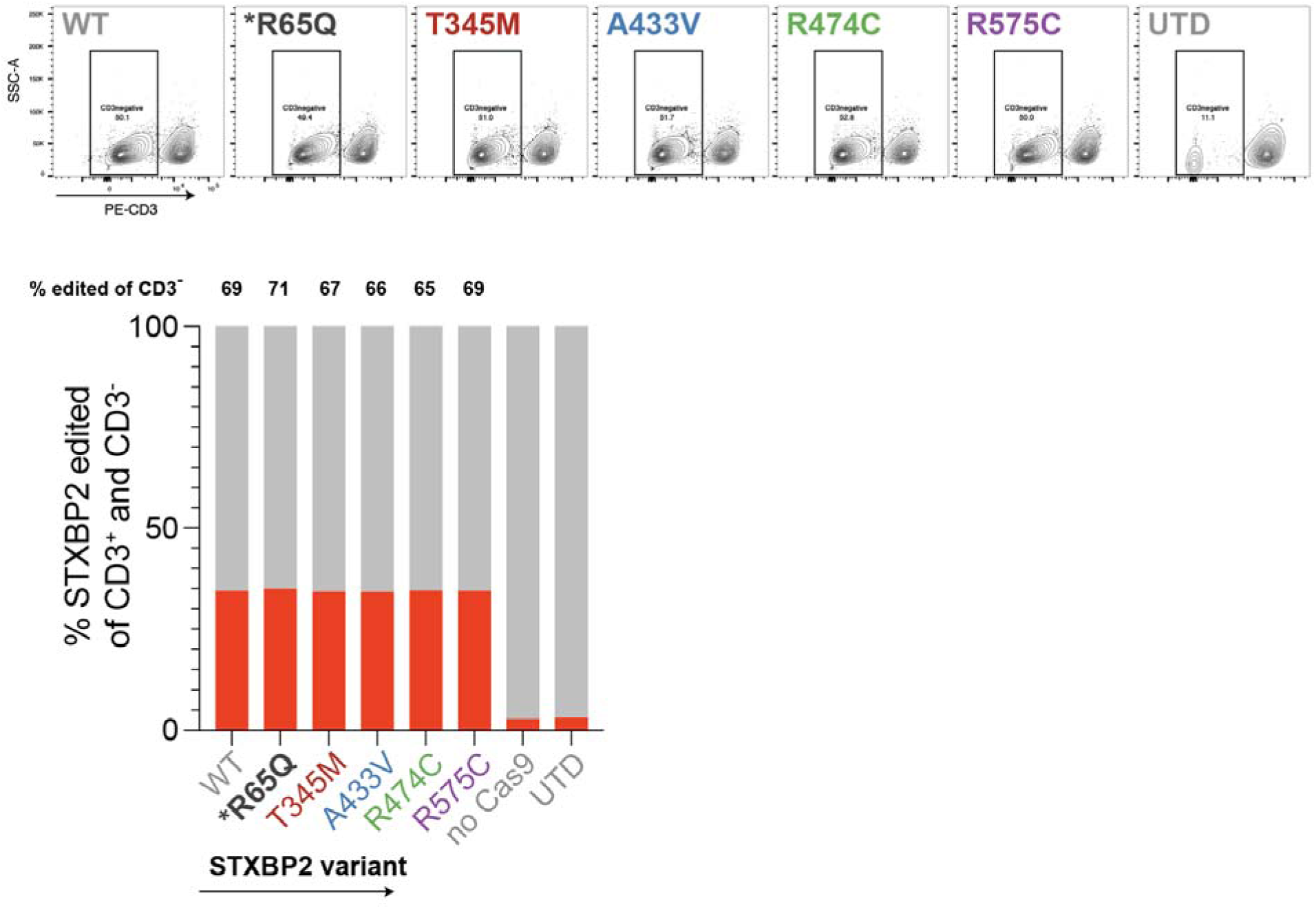
Knockout of wild-type STXBP2 in putative deleterious variant constructs. Primary CAR-T cells underwent transduction of the construct shown in Fig 2k. Followed by expansion and CD3 negative selection. All constructs were normalized to the same transduction efficiency (top) based on CD3 negativity. DNA was then extracted, and cells were sent for sequencing of an amplicon around the STXBP2 CRISPR-Cas9 knockout guide. Editing efficiency was quantified in CRISPResso after long-read sequencing through plasmidsaurus. Fraction of total STXBP2 knockout cells within the CD3-compartment is shown above each bar.

**Extended Data Figure 20.**
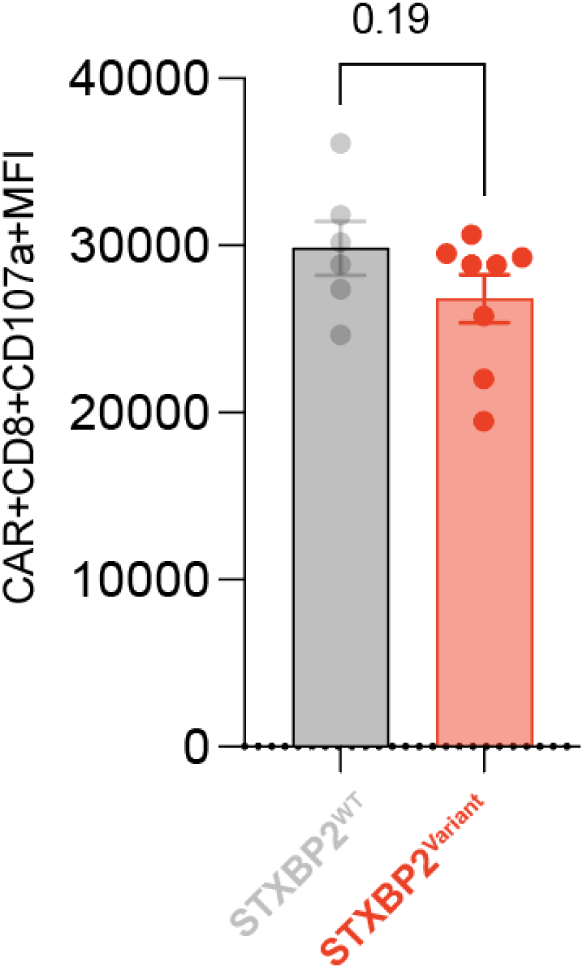
Degranulation of infusion product CAR-T cells from *STXBP2* variant carriers vs. non carriers in ZUMA-1. Infusion product CAR-T cells from patients were co-cultured with CD19-coated red blood cells for 4 hours. Degranulation was assessed via flow cytometry. Bars represent mean±SEM. P-value by unpaired t-test.

**Extended Data Figure 21.**
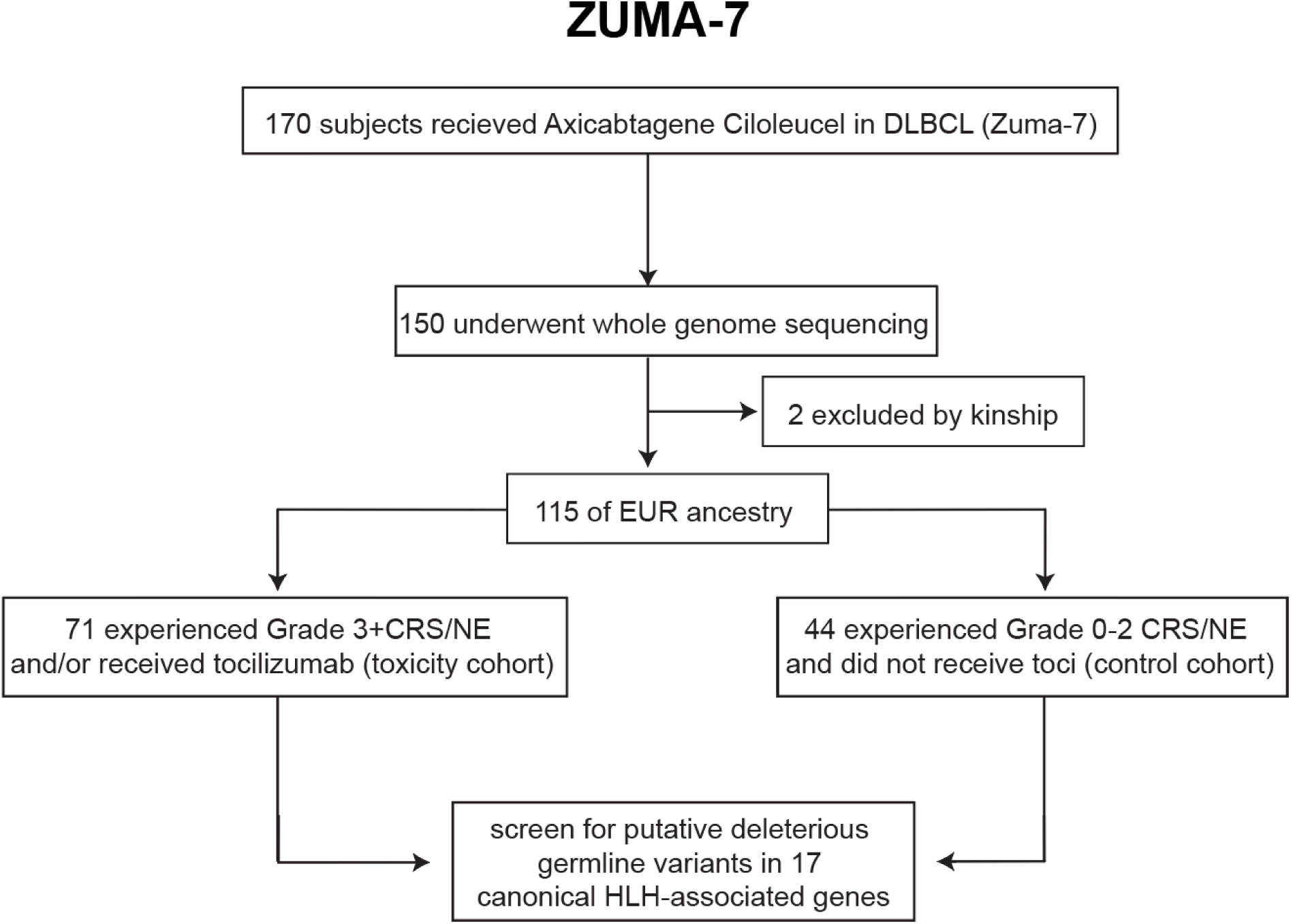
Diagram of ZUMA-7 subjects included for this study

**Extended Data Figure 22.**
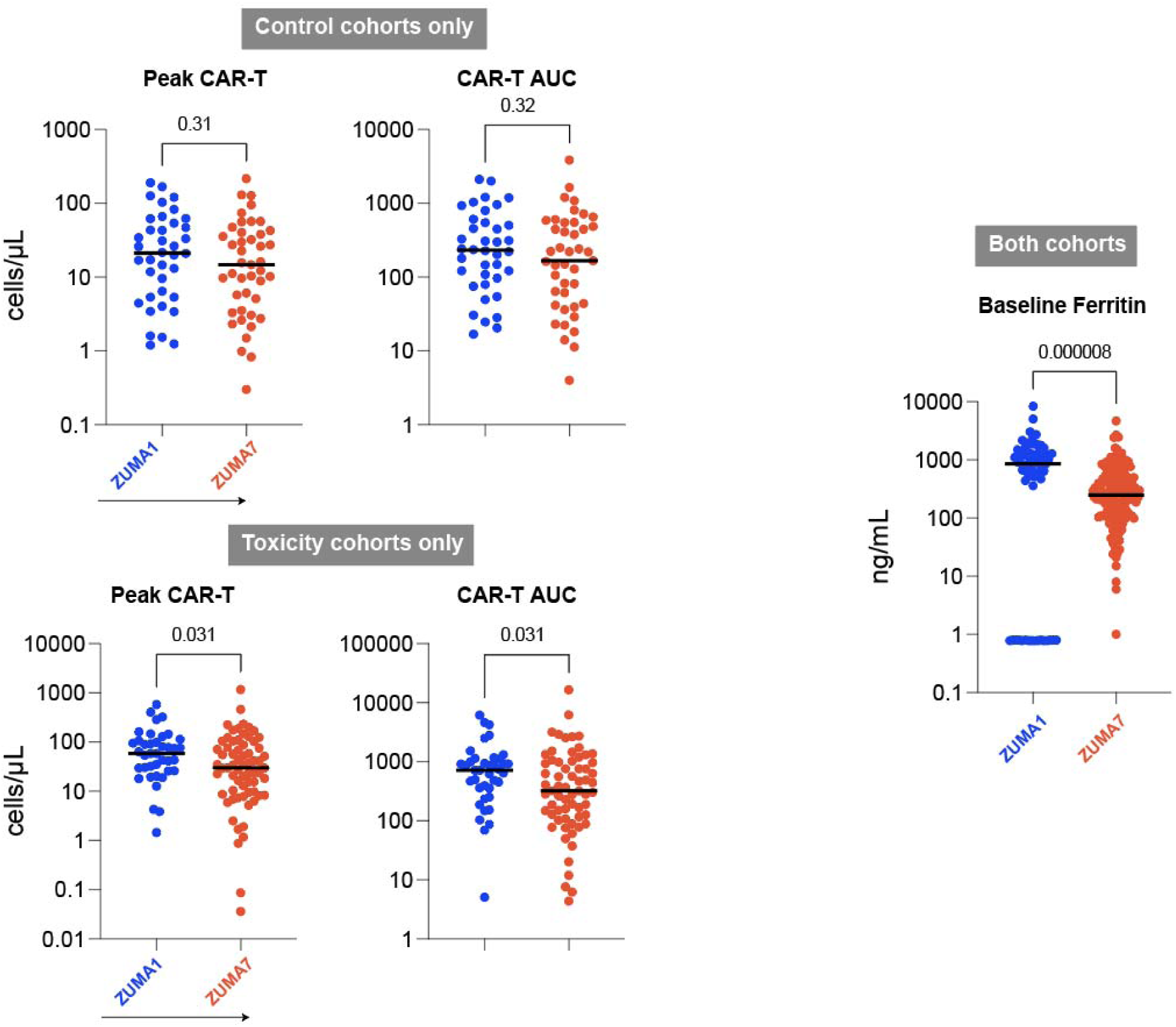
Peak and AUC CAR metrics from ZUMA-1 and ZUMA-7 control and toxicity cohorts, and baseline inflammatory state. Bars represent the median. P-values by Mann-Whitney U tests.

**Extended Data Figure 23.**
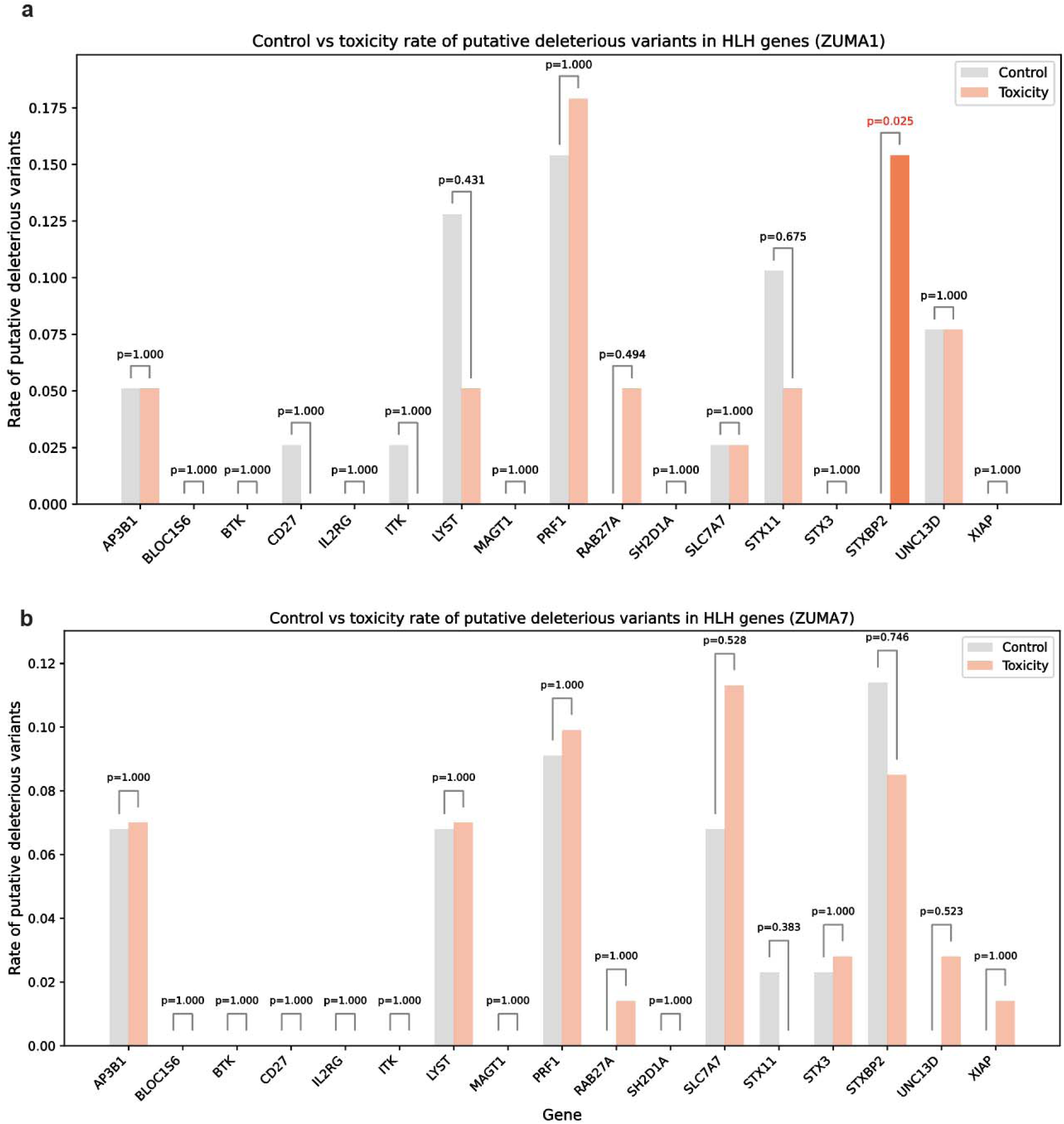
Rate of putative deleterious variants of HLH-associated genes in ZUMA1 and ZUMA7. P-values by Fisher’s Exact Test. a. ZUMA-1 cohort, b. ZUMA-7 cohort.

**Extended Data Figure 24.**
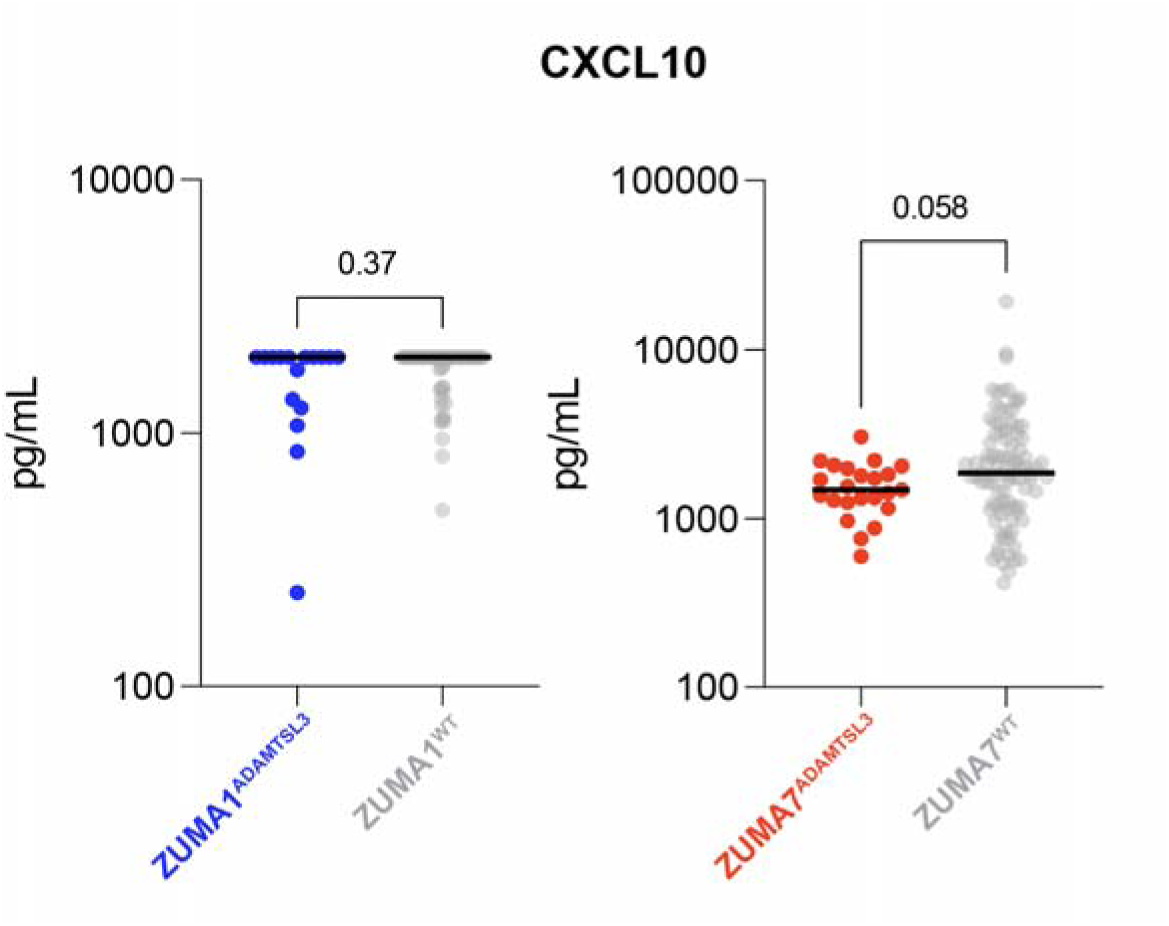
CXCL10 expression among ADAMTSL3 putative deleterious variant carriers versus wild-type patients in ZUMA1 and ZUMA7 Bars represent the median. P-values by Mann-Whitney U test.

**Extended Data Figure 25.**
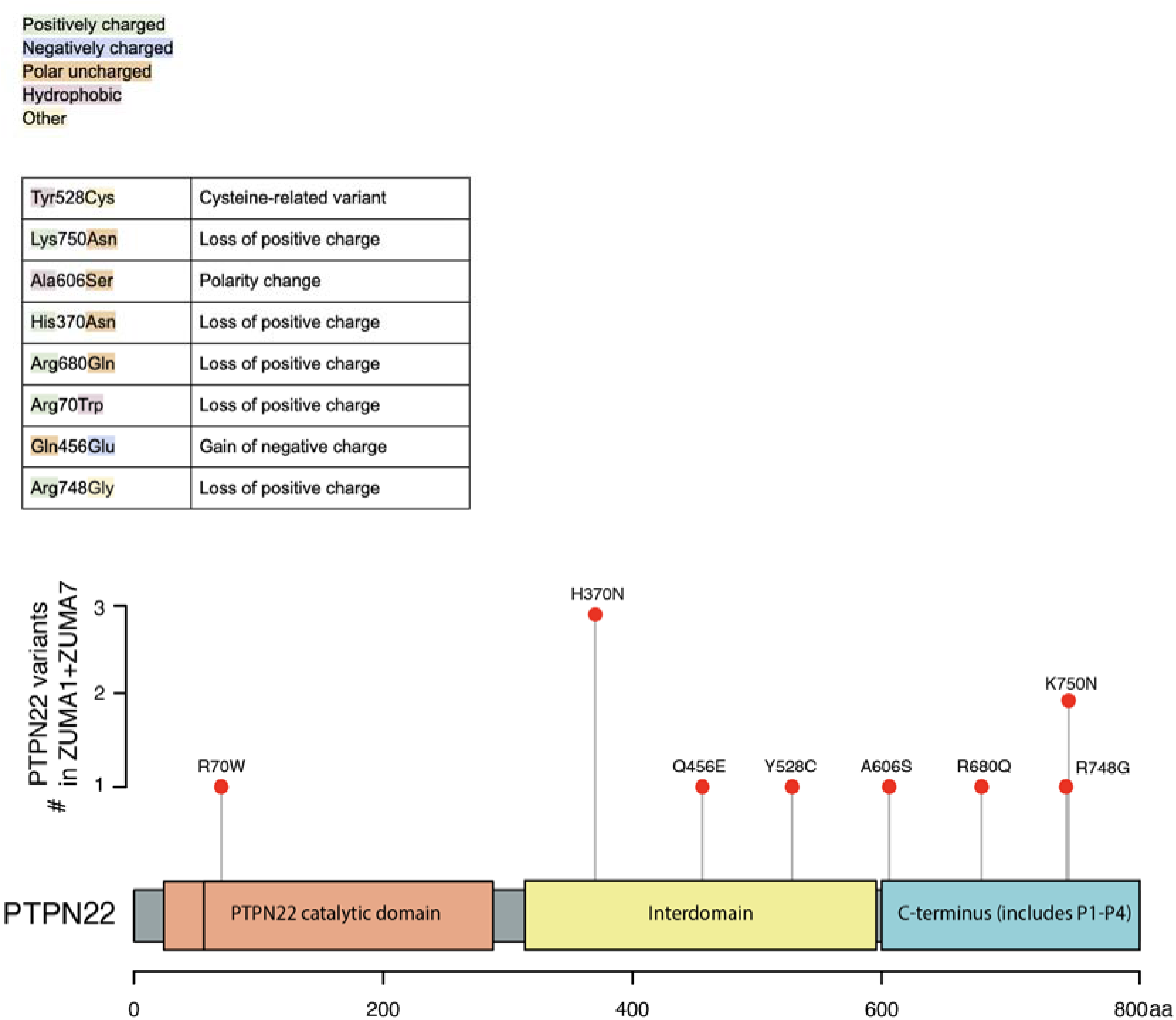
Consequences of Putative Deleterious Variants in PTPN22 among ZUMA1 and ZUMA7 patients

**Extended Data Figure 26.**
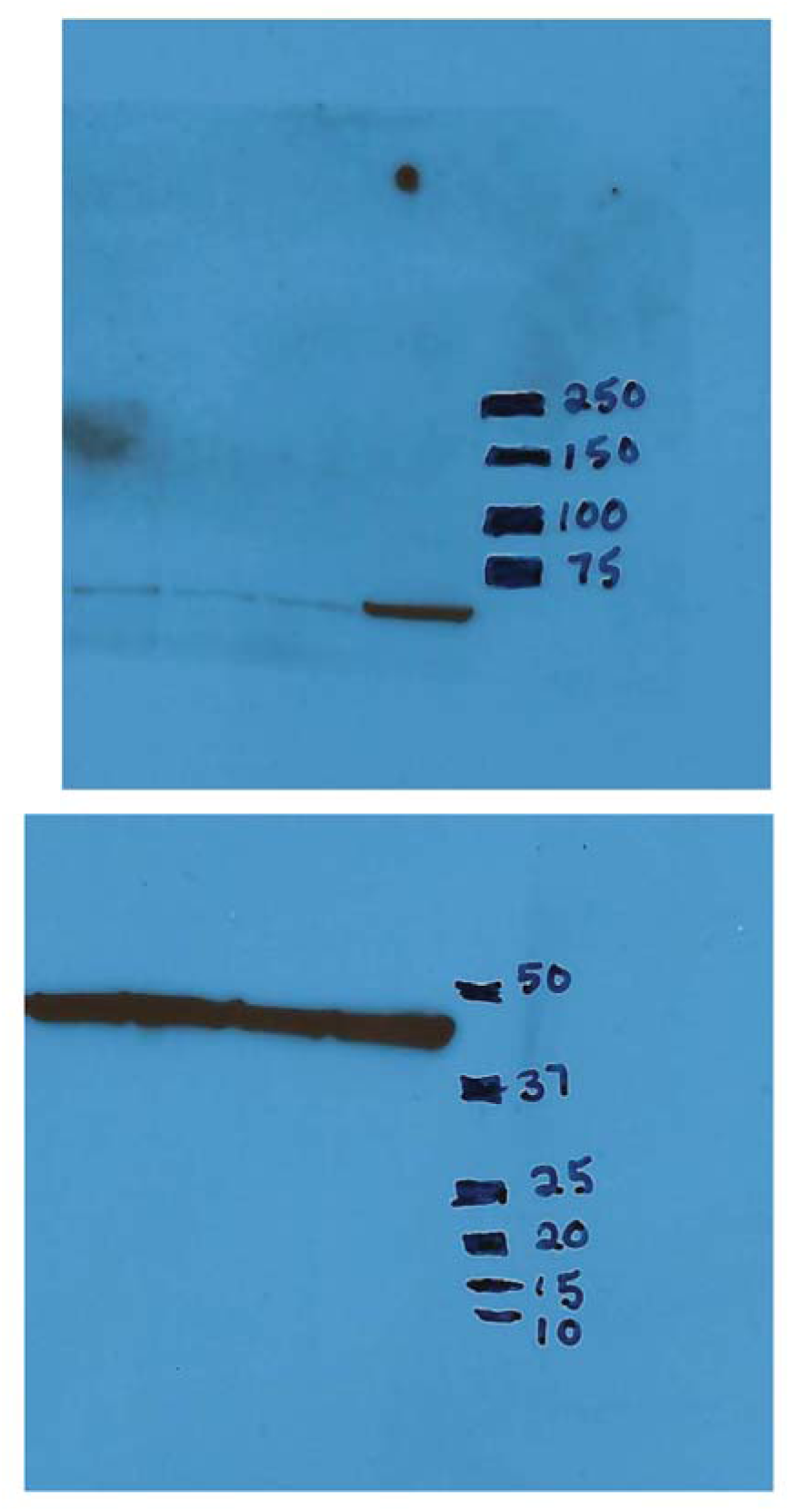
Full western blot for main text figure 1.

**Extended Data Table 1.**
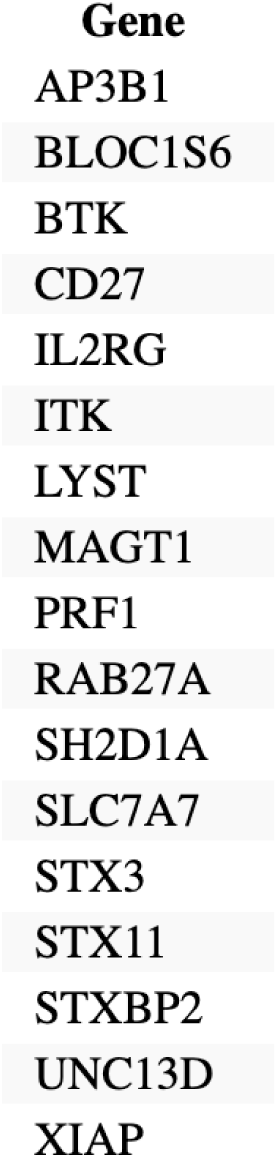
HLH-gene list. List of genes included in this study.

**Extended Data Table 2.**
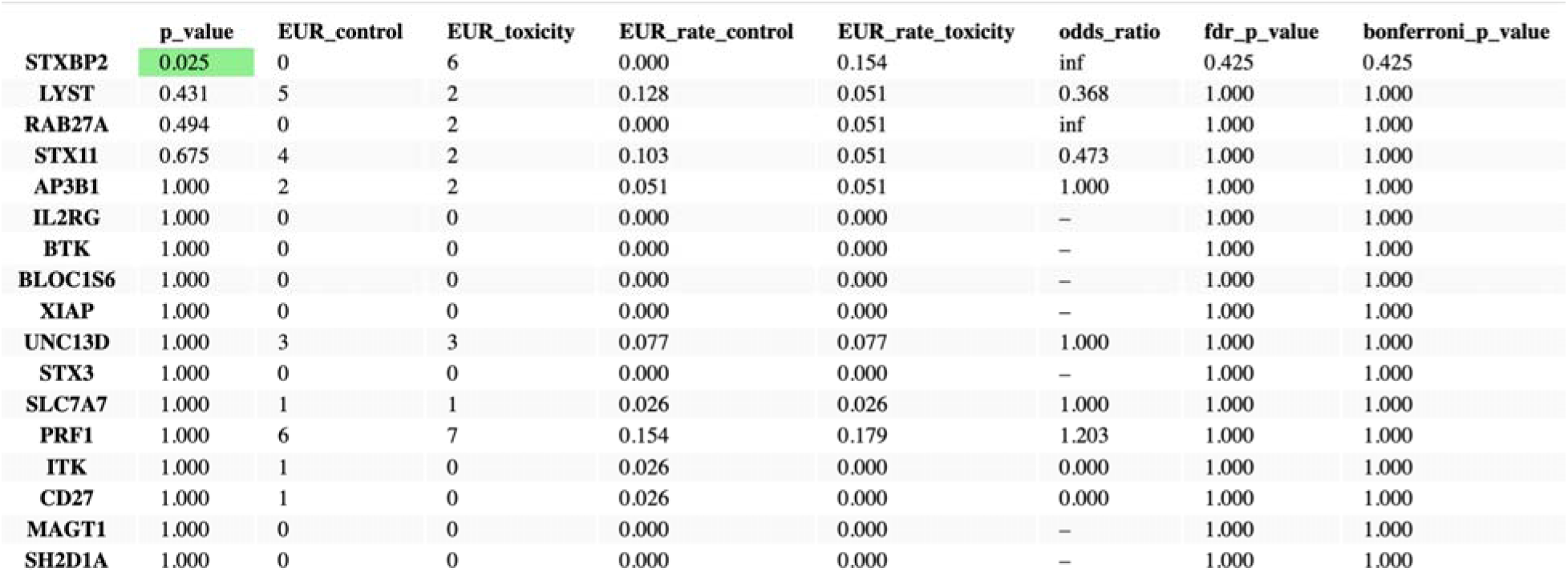
Gene-level burden of putative deleterious variants in HLH-associated genes. The percentage of each cohort with a given putative deleterious variant is shown. P-value and odds ratio calculated via Fisher’s Exact test. P-values adjusted for FDR using the Benjamini-Hochberg procedure and p-values adjusted using the Bonferroni correction are also shown.

**Extended Data Table 3.** Sequence Kernel Association Testing (SKAT) of *STXBP2* putative deleterious variants Given that a standard gene burden analysis relies on the assumption that all variants have the same effect size and direction, we leveraged SKAT, which is more robust when a fraction of variants could be noncausal (e.g., without strong *a priori* evidence of pathogenicity in the context of incomplete clinical databases). Similarly, our SKAT analysis showed an enrichment of putative deleterious germline variants in *STXBP2* in cases vs. controls (nominal p=0.0739 for ρ=1, p=0.0065 for optimal ρ).^96, 97^ Optimal rho is the value of rho to maximize statistical power, resulting in the best combination of SKAT and burden tests.

| $\rho$ | p-value |
| --- | --- |
| optimal | 0.0065 |
| 0 (burden) | 0.0051 |
| 1 (SKAT) | 0.0739 |

**Extended Data Table 4.** Total variants by classification stratified by gene Number of variants of any abundance according to class stratified by genes included in the study.

|  | AP3B1 | BLOC1S6 | BTK | CD27 | IL2RG | ITK | LYST | MAGT1 | PRF1 | RAB27A | SH2D1A | SLC7A7 | STX3 | STX11 | STXBP2 | UNC13D | XIAP |
| --- | --- | --- | --- | --- | --- | --- | --- | --- | --- | --- | --- | --- | --- | --- | --- | --- | --- |
| 3_prime_UTR_variant | 0 | 0 | 0 | 0 | 0 | 0 | 1 | 0 | 0 | 0 | 0 | 0 | 0 | 0 | 1 | 0 | 1 |
| inframe_deletion | 2 | 0 | 0 | 0 | 0 | 0 | 0 | 0 | 0 | 0 | 0 | 1 | 0 | 0 | 0 | 0 | 0 |
| intron_variant | 4 | 1 | 1 | 1 | 0 | 2 | 4 | 0 | 0 | 0 | 0 | 0 | 0 | 0 | 4 | 3 | 0 |
| missense_variant | 5 | 0 | 0 | 4 | 0 | 2 | 12 | 0 | 6 | 2 | 0 | 2 | 2 | 5 | 7 | 11 | 2 |
| splice_region_variant | 4 | 0 | 0 | 0 | 0 | 0 | 2 | 0 | 0 | 1 | 0 | 1 | 0 | 0 | 2 | 1 | 0 |
| synonymous_variant | 9 | 0 | 2 | 0 | 0 | 3 | 18 | 2 | 7 | 0 | 0 | 6 | 0 | 1 | 5 | 9 | 1 |

**Extended Data Table 5.** Common variant gene burden testing The percentage of each cohort with at least one common variant of each of the indicated genes is shown. P-value and odds ratio calculated via Fisher’s Exact test. P-values adjusted for FDR using the Benjamini-Hochberg procedure and p-values adjusted using the Bonferroni correction are also shown.

|  | p_value | EUR_control | EUR_toxicity | EUR_rate_control | EUR_rate_toxicity | odds_ratio | fdr_p_value | bonferroni_p_value |
| --- | --- | --- | --- | --- | --- | --- | --- | --- |
| STXBP2 | 0.377 | 34 | 30 | 0.872 | 0.769 | 0.490 | 1.000 | 1.000 |
| UNC13D | 0.472 | 28 | 24 | 0.718 | 0.615 | 0.629 | 1.000 | 1.000 |
| PRF1 | 0.737 | 33 | 35 | 0.846 | 0.897 | 1.591 | 1.000 | 1.000 |
| XIAP | 0.810 | 27 | 25 | 0.692 | 0.641 | 0.794 | 1.000 | 1.000 |
| AP3B1 | 1.000 | 39 | 39 | 1.000 | 1.000 | – | 1.000 | 1.000 |
| RAB27A | 1.000 | 0 | 0 | 0.000 | 0.000 | – | 1.000 | 1.000 |
| IL2RG | 1.000 | 0 | 0 | 0.000 | 0.000 | – | 1.000 | 1.000 |
| BLOC1S6 | 1.000 | 0 | 0 | 0.000 | 0.000 | – | 1.000 | 1.000 |
| STX11 | 1.000 | 1 | 1 | 0.026 | 0.026 | 1.000 | 1.000 | 1.000 |
| SLC7A7 | 1.000 | 39 | 39 | 1.000 | 1.000 | – | 1.000 | 1.000 |
| MAGT1 | 1.000 | 0 | 0 | 0.000 | 0.000 | – | 1.000 | 1.000 |
| LYST | 1.000 | 32 | 32 | 0.821 | 0.821 | 1.000 | 1.000 | 1.000 |
| ITK | 1.000 | 2 | 2 | 0.051 | 0.051 | 1.000 | 1.000 | 1.000 |
| CD27 | 1.000 | 39 | 39 | 1.000 | 1.000 | – | 1.000 | 1.000 |
| BTk | 1.000 | 9 | 8 | 0.231 | 0.205 | 0.860 | 1.000 | 1.000 |
| SH2D1A | 1.000 | 0 | 0 | 0.000 | 0.000 | – | 1.000 | 1.000 |
| STX3 | 1.000 | 0 | 0 | 0.000 | 0.000 | – | 1.000 | 1.000 |

**Extended Data Table 6.** Gene-level burden of rare synonymous germline variants in HLH-associated genes Percentage of each cohort with a rare variant of each of the indicated genes is shown. P-value and odds ratio calculated via Fisher’s exact test. P-values adjusted for FDR using the Benjamini-Hochberg procedure and p-values adjusted using the Bonferroni correction are also shown.

|  | p_value | EUR_control | EUR_toxicity | EUR_rate_control | EUR_rate_toxicity | odds_ratio | fdr_p_value | bonferroni_p_value |
| --- | --- | --- | --- | --- | --- | --- | --- | --- |
| AP3B1 | 1.000 | 2 | 1 | 0.051 | 0.026 | 0.487 | 1.000 | 1.000 |
| BLOC1S6 | 1.000 | 0 | 0 | 0.000 | 0.000 | – | 1.000 | 1.000 |
| BTK | 1.000 | 0 | 0 | 0.000 | 0.000 | – | 1.000 | 1.000 |
| CD27 | 1.000 | 0 | 0 | 0.000 | 0.000 | – | 1.000 | 1.000 |
| IL2RG | 1.000 | 0 | 0 | 0.000 | 0.000 | – | 1.000 | 1.000 |
| ITK | 1.000 | 2 | 1 | 0.051 | 0.026 | 0.487 | 1.000 | 1.000 |
| LYST | 1.000 | 1 | 1 | 0.026 | 0.026 | 1.000 | 1.000 | 1.000 |
| MAGT1 | 1.000 | 0 | 0 | 0.000 | 0.000 | – | 1.000 | 1.000 |
| PRF1 | 1.000 | 0 | 1 | 0.000 | 0.026 | inf | 1.000 | 1.000 |
| RAB27A | 1.000 | 0 | 0 | 0.000 | 0.000 | – | 1.000 | 1.000 |
| SH2D1A | 1.000 | 0 | 0 | 0.000 | 0.000 | – | 1.000 | 1.000 |
| SLC7A7 | 1.000 | 1 | 0 | 0.026 | 0.000 | 0.000 | 1.000 | 1.000 |
| STX11 | 1.000 | 0 | 0 | 0.000 | 0.000 | – | 1.000 | 1.000 |
| STX3 | 1.000 | 0 | 0 | 0.000 | 0.000 | – | 1.000 | 1.000 |
| STXBP2 | 1.000 | 1 | 0 | 0.026 | 0.000 | 0.000 | 1.000 | 1.000 |
| UNC13D | 1.000 | 1 | 0 | 0.026 | 0.000 | 0.000 | 1.000 | 1.000 |
| XIAP | 1.000 | 0 | 1 | 0.000 | 0.026 | inf | 1.000 | 1.000 |

**Extended Data Table 7.**
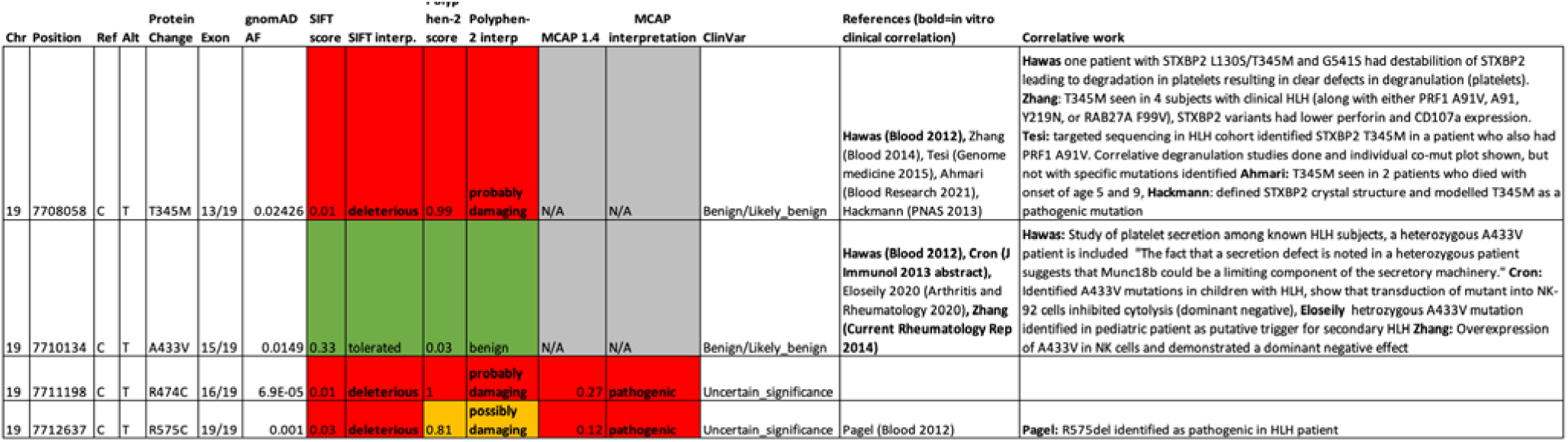
Individual variant characteristics and literature references/summary. References highlighted in bold represent studies in which *in vitro* experiments were performed to assess variant function. Chr-chromosome, Ref-reference, alt-alternate, AF-allele frequency, SIFT-sorting intolerant from tolerant, Polyphen-Polymorphism Phenotyping v2, MCAP-mendelian clinically applicable pathogenicity. N/A not available. M-CAP Scores are only available where no superpopulation has minor allele frequency above 1%. In the correlative work column, the first author of the cited reference is listed

**Extended Data Table 8.** Clinical characteristics of individuals by *STXBP2* putative deleterious variant Cohort means are shown below for CAR-T cell peak and AUC expansion.

|  | Subject ID | Age | Inferred Ancestry | Sex | CAR peak (cells/uL) | CAR AUC | Time to peak (days) | Neurotoxicity | CRS | Corticosteroids use | Tocilizumab use | Response | Best Response | Last Response |
| --- | --- | --- | --- | --- | --- | --- | --- | --- | --- | --- | --- | --- | --- | --- |
| **19:7708058 (T345M)** |  |  |  |  |  |  |  |  |  |  |  |  |  |  |
| MGH 1028 VAF 0.3886 C/T | 1 | <65 Years | EUR | Male | 20 | 149 | 8 | G1/2 | G1/2 | Y | Y | Responder | PR | PR |
| MGH 1044 0.36 C/T | 2 | >=65 Years | EUR | Female | 43 | 621 | 8 | G3+ | G1/2 | Y | Y | Responder | CR | CR |
| **19:7710134 (A433V)** |  |  |  |  |  |  |  |  |  |  |  |  |  |  |
| MGH 16-591 0.41 C/T 3+NTX | 3 | <65 Years | EUR | Male | 75 | 754 | 8 | G3+ | G1/2 | N | Y | Responder | CR | CR |
| MGH 16-597 .48 C/T 3+NTX | 4 | >=65 Years | EUR | Male | 78 | 934 | 8 | G3+ | G1/2 | Y | Y | Responder | CR | CR |
| **19:7711198 (R474C)** |  |  |  |  |  |  |  |  |  |  |  |  |  |  |
| MGH 16-574 0.48 C/T | 5 | <65 Years | EUR | Male | 30 | 103 | 8 | G1/2 | G3+ | Y | Y | Non-responder | PD | SD+PD |
| **19:7712637 (R575C)** |  |  |  |  |  |  |  |  |  |  |  |  |  |  |
| MGH 16-577 0.48 C/T 3+NTX | 6 | >=65 Years | EUR | Male | 19 | 236 | 8 | G3+ | G1/2 | Y | Y | Responder | CR | CR |

|  | cohort median |  |  |
| --- | --- | --- | --- |
| controls | 21 | 238 | 8 |
| toxicity | 59 | 733 | 8 |

## Notes

### Author Declarations

We obtained available genomic DNA from patients in the ZUMA-1 and ZUMA-7 clinical trials. All patients had provided written informed consent as part of the ZUMA-1 and ZUMA-7 studies at their respective institutions. None of the authors of this manuscript had access to identifiable patient information and nor did they participate in the consent process for the individuals. For primary T-cells used in experiments: Human T cells were purified (Stem Cell Technologies, #15061) from healthy donor leukopaks whose use was determined to be non-human subjects research by the Institutional Review Board (IRB) at the Massachusetts General Hospital (MGH).

