## Supplemental appendix for "Genomic correlates of clinical CAR-T cell activity"

**Supplementary Information**

**Germline Variant Calling from the sequencing data**

Germline variants were called from the whole genome sequencing BAM files using DeepVariant (version 0.8.0, docker: gcr.io/deepvariant-docker/deepvariant:0.8.0), a deep-learning based variant calling tool that had demonstrated superior sensitivity and specificity than GATK based joint-genotyping.^1,2^ Germline variants with Genotype Quality score above 20 and “PASS” in the FILTER column of the variant call format (VCF) files were chosen, and this final set of high-quality variants was combined into cohort-level VCF files using GATK (version 3.7) tool “CombineVariants”. Finally, the “vt” tool (version 3.13) was used on the cohort-level VCF files to normalize and decompose multiallelic variants. To evaluate insertion/deletion (Indel) distribution and the ratio of transitions to transversions we used “bcftools” (version 1.9) tools; “stats” and “plot-vcfstats”.

**Relatedness Analysis**

We conducted a genetic relatedness analysis using the cohort-level VCF files in two steps. First, we used GENESIS (version 2.12.0) tool “PC-AiR” to conduct a principal components analysis (PCA) using identified germline variants for the detection of population structure in the toxicity and control cohort. We then used the GENESIS tool “PC-Relate”^7^ implemented in “Hail” (version 0.2.11, https://github.com/hail-is/hail) to estimate kinship coefficients for every possible pair within a cohort. In ZUMA-1, only one sample pair had a kinship coefficient above 0.125 (0.157) indicating a genetic relatedness of second-degree relatives; however, this sample was already excluded from the burden analysis during quality control from ancestry inference. In ZUMA-7, three sample pairs had kinship coefficients above 0.125 (0.473, 0.170, 0.137). Two samples were removed such that no second-degree relatives remained, one of which was excluded previously during quality control from ancestry inference.

**Functional and Clinical Annotation and Prioritization of Germline Variants**

Germline variants in the cohort-level VCF files were annotated using Variant Effect Predictor (VEP, version 110, GRCh38)^8^ with dbNSFP (version 4.9a, GRCh38)^9^ and ClinVar (Release Dec2024, GRCh38) plug-ins.^10^ Pymol version 2.5.2 was used to model predicted effects of reported variants. Model 4CCA (https://www.rcsb.org/structure/4cca) deposited by the authors of the manuscript reporting the crystal structure for STXBP2 was used in Pymol.^11^

**Cancer-Free Control Cohort**

WES BAMs were aligned to GRCh37 from a total of 24,128 adult unrelated individuals without known cancer diagnosis. They were collected from the following studies: Autism Sequencing Consortium (ASC) (Database of Genotypes and Phenotypes (dbGAP):phs000298.v4.p3), Framingham Cohort (dbGAP:phs000007.v32.p1), Multi-Ethnic Study of Atherosclerosis (MESA) Cohort (dbGAP: phs000209.v13.p3), National Heart, Lung and Blood Institute (NHLBI) GO-ESP: Lung Cohorts Exome Sequencing Project (dbGAP: phs000291.v2.p1), 1000 Genomes Project9. All control samples were processed using methods identical to those used for the discovery cohort except for the following: the discovery cohort utilized VEP version 110 and ClinVar release Dec2024, and the cancer-free control cohort used VEP version 104.3 and ClinVar release of Aug2022.

**CAR design**

CAR constructs were synthesized and cloned into a second-generation lentiviral backbone behind a human EF-1a promoter (Genescript). Our anti-CD19 construct utilized a CD28 costimulatory domain, similar in design to axicabtagene ciloleucel. RNA guide sequences for TRAC (AGAGTCTCTCAGCTGGTACA) and STXBP2 (ACACATCTGCGATATGCATG, CGGCCTAGCTCACTGAACAG, GTCCAGCAGATCATACGCCA) drawn from the Brunello guide library^12^. The control scramble guide has been used previously (GGTTCTTGACTACCGTAATT^13^). All constructs contained a transgene encoding the fluorescent reporter mCherry to evaluate transduction.

**CAR-T cell production**

Human T cells were purified (Stem Cell Technologies, #15061) from healthy donor leukopaks whose use was determined to be ‘non-human subjects research’ by the Institutional Review Board (IRB) at the Massachusetts General Hospital (MGH). These human T cells were activated using CD3/CD28 Dynabeads (1 T cell to 3 beads) (ThermoFisher Scientific, 40203D) on Day 0, followed by transduction with a lentiviral vector encoding the CAR on day 1 (24 h later). T cells were cultured in R10 media with 20 IU/mL of recombinant human IL-2, penicillin, and streptomycin. T cells were resuspended at 5x10^6^/100uL in Opti-MEM (thermo fisher scientific) after debeading on day 5 and electroporated with 10ug Cas9 mRNA (triLink) at 360V x .001 ms. 3-5 days later CD3- T cells were isolated via positive column selection (EasySep Human APC Positive Selection Kit II; StemCELL Technologies). Deletion of CD3 was confirmed by flow cytometry and STXBP2 by western blot. Cell transduction efficiency ranged from 92-95%.

**Western Blot**

Ten micrograms of whole cell lysates from primary CAR-T cells were separated by SDS-PAGE on 4-12% Bis-Tris gels. Gels were transferred using the Novex iBlot2 Gel Transfer Device (invitrogen) and iBlot 2 Nitrocellulose Transfer stacks (Invitrogen) according to manufacturer’s protocols. Gels were transferred using a 7-minute dry transfer protocol (1 minute at 20V, 4 minutes at 23V, 2 minutes at 25V). Blots were blocked using 5% nonfat dry milk (Bio-Rad) in 1X Tris-Buffered Saline, 0.1% Tween® 20 Detergent (TBST) (Santa Cruz Biotechnology) for at least 1 hour at 25C. The membrane was then cut into two pieces, using the protein ladder as a guide to separate between the expected STXBP2 protein band, and the B-actin protein band. The top membrane was washed once in TBST and probed overnight at 4C using anti-STXBP2 mouse monoclonal antibody (Proteintech Cat#66238) at a dilution of 1:5,000 in 5% BSA (invitrogen). The bottom membrane was washed once in TBST and probed overnight at 4C using anti-B-actin rabbit monoclonal antibody (Cell Signaling Technology #4970) at a dilution of 1:12,000. Membranes were washed three times for 10 min with TBST and incubated for 1 hour with either Mouse IgG HRP Linked Whole Antibody (GE Healthcare) (top membrane) or Goat anti-Rabbit IgG (H+L) Secondary Antibody HRP (Invitrogen #31460) (bottom membrane) diluted in 5% non-fat dry milk TBST for one hour at room temperature at the indicated dilutions (1:15,000 and 1:30,000 respectively). Membranes were then washed three times for 10 min each with TBST And developed with Signalfire ECL Reagent (CST).

**CAR-T Degranulation assay**

1x10^5^ CAR-T cells were co-cultured with 1x10^5^ CD19-expressing K562 cells for 4 hours in 96-well plates after brief centrifugation. Cells were resuspended in 2% FCS (FACS buffer) and stained for CD107a (BD Biosciences APC clone H4A3) for 30 minutes at 4C. CD71 (Biolegend FITC), CD45 (Biolegend PECy7 clone HI30), CD3 (BDBiosciences APC-H7 clone SK7), CD4 (BD Biosciences V450 clone SK3), and CD8 (BD Biosciences APC R700 Clone SK1) were then added for another 15 minutes. After washing cells were stained in fixable viability dye 575V (BD Biosciences FVS575V) for 5 minutes at 37C. Following washing, cells were fixed and permeabilized followed by staining for intracellular IFNγ (Biolegend BV785 Clone 4S.B3) for 30 minutes. After additional washes cells were analyzed on a BD FACS lyric. All flow cytometric data was analyzed in Flowjo (version 10.8.1). Reported percentages represent the K562-stimulated condition minus the unstimulated condition. Individual replicates and *STXBP2* guide knockouts were averaged and compared to controls using statistical tests as described in figure legends.

**Myeloid cell co-culture**

Monocytes were purified with EasySep™ Human Monocyte Isolation Kit (Stemcell Technologies 19359) from healthy donor PBMC. Isolated monocytes were then plated and differentiated into GM-CSF macrophage subsets as described previously.^14^ In brief, monocytes were cultured in the presence of recombinant GM-CSF (Biolegend, 5ng/mL) for 7 days in the bottom of a transwell plate at 1x10^5^ cells/well in a 48-well plate and left in this original position (not transferred). In the top of the transwell insert CAR-T cells were co-cultured with Nalm6 tumor cells at a 1:1 effector-to-target ratio. Supernatant was collected at 72 hours and frozen at -80C for further analysis.

**Real Time Cytotoxicity assay**

Plates were coated with anti-CD71 (Biolegend clone: CY1G4, unconjugated) capture antibody overnight followed by tumor seeding (1x10^5^/well of GFP expressing Jeko, Nalm6, and CD19-K562). CAR-T cells were added at the indicated ratios followed by monitoring on an incucyte SX5.

**CD19-coated red blood cell stimulation assay**

CD19-coated RBCs were plated in 96-well plates with three different CD19 surface densities in separate wells (100,000 cells/well) and co-cultured with Axi-cel products (100,000 cells/well). Uncoated RBCs served as negative controls. 5μl of PE-CD107a antibody (BioLegend cat no. 328608) and 1μl of Golgi Stop (BD Bioscience cat no. 555029) were added to each well. After 4 hours of co-culture, cells were washed with cold PBS and fixed with PFA (BD Bioscience cat no. 555028) for 15 minutes at 4°C. Then, cells were stained for extracellular markers: PE-Cy7-CD3 (BioLegend cat no. 317334), BV510-CD8 (BioLegend cat no. 344732), AF488-anti-FMC63 (internal antibody) and Live/Dead (LIVE/DEAD Fixable Near-IR Dead Cell Stain Kit, Thermo Fisher Scientific cat no. L34976), followed by permeabilization and intracellular staining of IFN-γ (AF647-IFN-γ; cat no. 502516). Flow cytometric data was acquired using a BD LSRFortessa and analyzed with FlowJo software.

**Luciferase Killing Assay**

CAR T cells normalized with UTDs to a transduction efficiency of 49% were resuspended at 1e6 cells/mL. In a 96 well flat-bottomed plate, CAR T cells were plated in triplicate at a 10:1 E:T ratio in row A and diluted 1:3 down rows B-G. 1e4 Nalm6 BID KO or Nalm6 WT cells were plated to their corresponding wells. The co-cultures were incubated overnight at 37°C for 18 hours. The next morning, the plates acclimated to room temperature and were then spun down at 1800rpm for 5 minutes. The supernatant was dumped and 50 μL of R10 followed by 50 μL of Bio-Glo Reagent were added carefully into each well (Promega, G7940). Each plate was incubated at room temperature for 15 minutes before measuring luminescence on the Spectramax luminometer.

**DNA isolation and sequencing**

gDNA was extracted and purified from a minimum of 1e6 cells using the DNeasy Blood and Tissue Kit (Qiagen, 69504). Resulting gDNA was stored at -20°C until PCR amplification. gDNA was amplified with PCR primers that were chosen using the tool primer-BLAST and Q5 High Fidelity 2x Master Mix reagent (NEB, M0492L). PCR was conducted following the protocol associated with the Q5 High Fidelity 2x Master Mix reagent on New England Biolabs website. PCR products were mixed with 6x loading dye (NEB, B7024S) and loaded onto a 1% agarose gel. SYBR safe was added at a ratio of 1:10000 for DNA visualization (Thermo, S33102). The gel was run for 45 minutes at 115V and then imaged. Bands at the proper product length were excised and the DNA was extracted using the Nucleospin Gel and PCR kit (Macherey-Nagel, 740609.25). After elution, the DNA concentration in the eluate was measured by nanodrop. Following Quintara guidelines, the purified DNA was resuspended at a concentration depending on product length and primer was added. Samples were sent to Quintara for sanger sequencing and then analyzed EditR or sent to Plasmidsaurus for nanopore sequencing and analyzed on CRISPResso2.

**Cytokine Analysis**

Frozen supernatant cytokines were analyzed using the Human Inflammatory Panel 20-plex ProcartaPlex Panel (Thermo Fisher Scientific) and run on a FLEXMAP 3D System (Luminex).

Supplemental Information References
